# Calibration-derived decoder discriminability is associated with online P300-speller accuracy, but the fitted mapping does not transport across cohorts

**DOI:** 10.64898/2026.07.30.26359353

**Authors:** Alon Gorenshtein, Yosef Adiniaev, Tom Liba, Yiftach Barash, Eyal Klang, Oved Daniel

## Abstract

**Objective:** A calibration-derived score has been related to online P300-speller accuracy in the same session, always within the cohort measured. We evaluated whether a fitted mapping from that score to expected accuracy transports to withheld cohorts.

**Approach:** Retrospective secondary analysis of BigP3BCI 1.0.0, the largest published evaluation of this relationship to date: 18 of 20 source studies yielded eligible online outcomes, contributing 271 participants, 739 session-condition records, and 19,611 character selections. Four cohorts document an amyotrophic lateral sclerosis (ALS) population. The predictor was calibration-derived decoder discriminability: cross-validated discriminability of a classifier fitted only to calibration epochs. One source study at a time was withheld from development. Between-cohort variation was summarised by the random-effects standard deviation tau, with participant-clustered standard errors.

**Main Results:** The association was positive in all 18 cohorts but varied widely in magnitude and precision (Pearson r 0.190 to 0.928; participant-level r = 0.714, p < 0.001). Pooled estimation error was 0.098 (95% CI 0.091 to 0.107) against a benchmark of 0.146. Calibration did not transport: the intercept had tau 0.87, with a 95% interval for an unrepresented cohort of -1.97 to 1.85 on the log-odds scale, and the slope varied more than tenfold across cohorts (tau 0.43, unrepresented-cohort interval 0.111 to 2.005). A protocol proxy for the stopping rule, median time per selection, reduced the between-cohort slope variance by 66%. The four ALS cohorts alone gave an uncorrected between-cohort slope spread of 0.223, against 0.560 overall.

**Significance:** The score carries a reproducible signal about accuracy in the corresponding session, but the mapping between them is cohort-specific: usable for ranking sessions within a setting, not for reporting expected accuracy elsewhere without local recalibration. Few-cohort evaluation, as the ALS subgroup illustrates, understates how much performance varies elsewhere.

## Introduction

The P300 speller lets a person select characters from a visual matrix using event-related potentials rather than movement, one of the few communication routes available when motor control is lost.[1,2] It has been evaluated repeatedly in amyotrophic lateral sclerosis (ALS), including long-term independent home use,[3–6] and sits alongside implanted systems among the communication options for severe paralysis.[7,8]

Online spelling accuracy varies widely between users and sessions, and some users never reach usable accuracy.[9] This motivated work asking whether performance can be anticipated before a session begins: neurophysiological features from rest or a calibration block relate to later brain-computer interface performance,[10] including P300-speller accuracy in ALS specifically,[11,12] and attentional and motivational state have also been linked to performance.[13,14] Mainsah and colleagues went furthest, deriving P300-speller accuracy analytically from a calibration-derived detectability index, to estimate performance without extensive online testing.[15] A projected-accuracy measure and a separate multi-feature predictor have made related within-session or within-study predictions.[37,38]

Two features of that literature limit what it can support. First, the relationships were established within the cohort measured; this does not establish that a mapping fitted in one set of cohorts will estimate accuracy in a cohort it has never seen, which is what matters if a score is to be used anywhere other than where it was developed. Second, these studies report discrimination, usually a correlation or an area under the receiver operating characteristic curve, which describes whether users can be ranked but not whether the estimated accuracy is numerically close to the accuracy that follows. To our knowledge, no published study has evaluated whether a calibration-derived score’s fitted mapping to online accuracy transports to a cohort withheld from its development, as distinct from its within-cohort association or discrimination.

Evaluating transportability also requires more cohorts than this literature has used: the cohort is the unit of replication, and the spread of performance across withheld cohorts determines the interval a reader should expect in their own setting.[16,17] Public archives that aggregate many P300-speller studies under a shared montage now make more withheld cohorts available.[18] A recent cross-dataset evaluation aligned P300 signals across archives to improve classification transfer, a different question from whether a fitted calibration-to-accuracy mapping itself transports numerically.[39]

We evaluated whether a calibration-derived score estimates online session accuracy in the corresponding session, in cohorts withheld from model development, across every source study in a public archive that yields eligible online outcomes. The score, referred to throughout as calibration-derived decoder discriminability to keep it distinct from a physiological marker of user aptitude, is the cross-validated discriminability of a classifier fitted to a session’s calibration epochs. The ALS cohorts were the originally planned subgroup before the design widened (Methods). Because the archive also contains cohorts without a documented ALS population, we further asked whether a mapping developed without any ALS data transports to the ALS cohorts, and whether the calibration-to-accuracy relationship itself differs between cohort types.

## Methods

### Study Design and Reporting

This was a retrospective secondary analysis of a public archive of legacy online P300-speller recordings, using an internal-external cross-validation across source studies: an entire source study was withheld from model development and the fitted mapping evaluated in that withheld cohort.[16,17] Every cohort comes from one harmonised archive rather than a separately assembled dataset, so this is not an external validation in the strict sense, but what is withheld, the source study, is what the transportability question is asked about. Reporting follows the TRIPOD statement.[19]

No analysis plan was registered. The four ALS cohorts were the originally planned subgroup before the design was widened; the sensitivity and comparator analyses were listed before either was run on the widened cohort but after the primary analysis had been fitted, and the protocol-descriptor moderator analysis was added later still, so none is prespecified in the sense a registered plan would establish (the repository’s commit history records these dates).

Calibration and online blocks come from the same recording session throughout. Every timestamp is de-identified, so the separation enforced here is between protocol phases, not demonstrated temporal precedence; an analysis using a preceding session’s calibration recording is reported separately, resting on the assumption that lexical session-identifier order matches recording order, which de-identified timestamps cannot confirm.

### Data Source and Cohort

The source was BigP3BCI version 1.0.0.[18] The downloaded archive was fixed by SHA256 digest (eea294aa34e9ed11e5a25d07e30aeefdf8b2d467a8309e2c38405a289afcd72f) before ingestion, and every selected European Data Format file was checked against the distributor checksum manifest.

All 20 documented source studies were screened; every study supplied the shared 16-channel montage at 256 Hz, so montage compatibility excluded none. Two studies contributed no eligible online outcome, for the archive-level reasons given in Table 1, and are retained in the study inventory since their exclusion follows from the archive rather than any analysis choice. The archive documentation identifies an ALS study population for four source studies; the remainder are described as other cohorts rather than healthy or control cohorts, since the documentation does not support that characterisation. Participant identifiers are study-scoped; because the source studies draw from a limited number of contributing laboratories, participant overlap across studies cannot be ruled out from the archive’s documentation (supplement, S1).

**Table 1.**
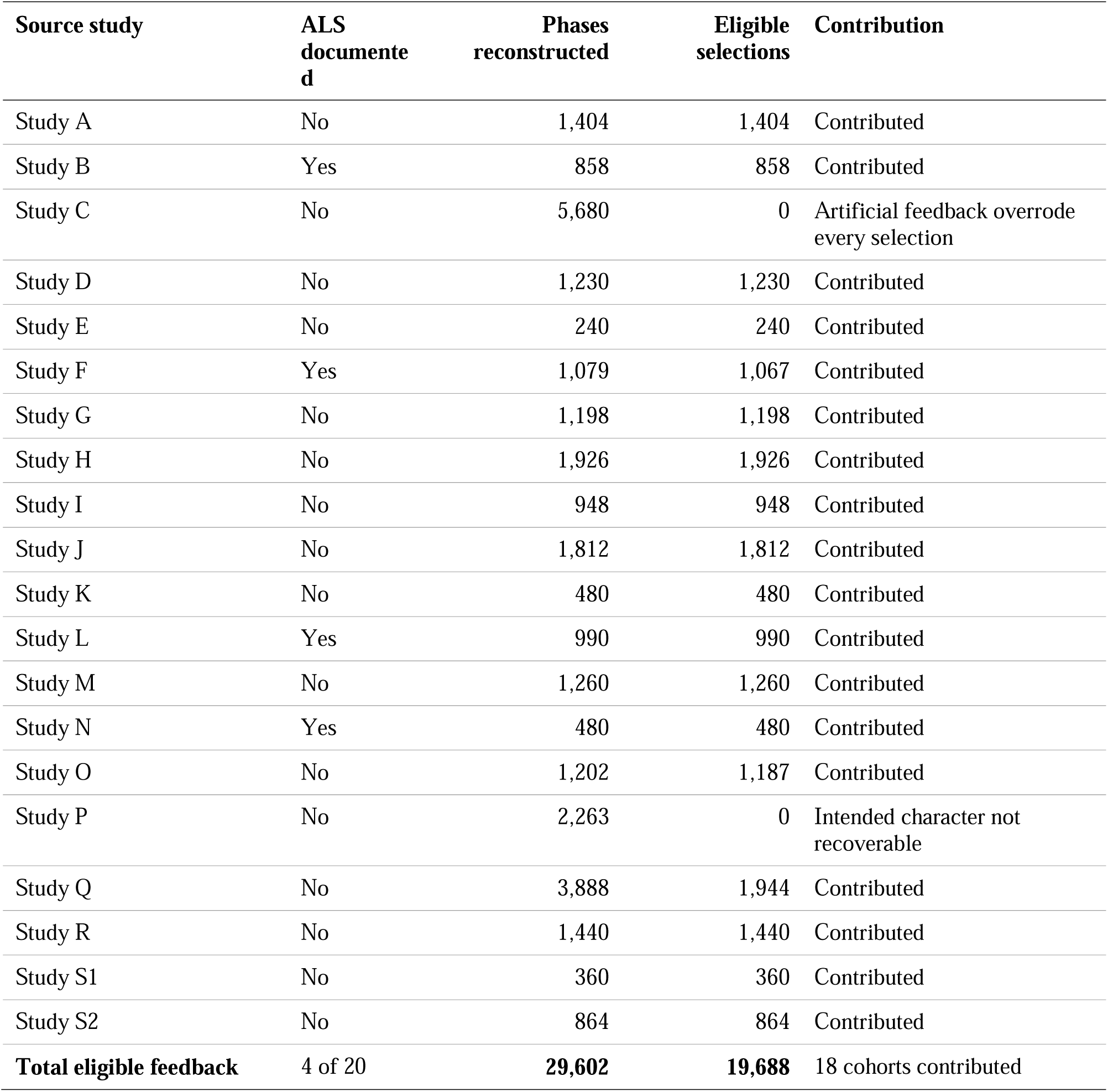
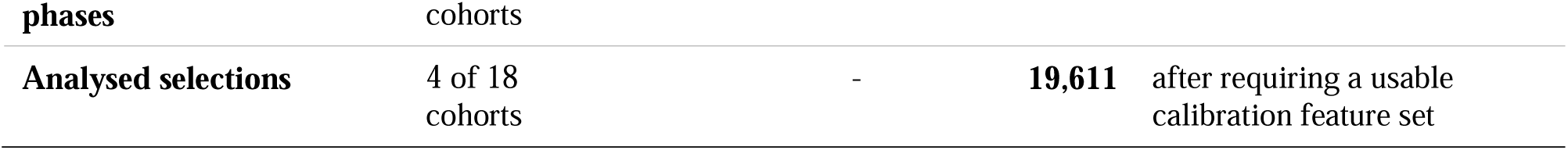
Source studies screened, and their contribution to the analytic set. All 20 documented source studies supplied the shared 16-channel montage at 256 Hz. Two contributed no eligible online outcome: Study C, in which artificial feedback overrode the selection in all 5,680 reconstructed feedback phases, and Study P, in which the intended character could not be recovered in any of 2,263 phases. Cohorts are marked according to whether the archive documentation identifies an ALS study population. The two totals in the final rows differ by 77 selections, all of them in Study B: two sessions of one Study B participant were archived with Test-phase recordings only and carry no calibration block at all, so no calibration score exists for them and their 38 and 39 selections cannot enter a model that takes that score as its predictor. Every other eligible selection in the archive is analysed, and the per-cohort selection counts in Table 2 are the analysed counts.

### Calibration Predictor

Only Train-phase files informed the predictor. A calibration event was a rising StimulusBegin transition during phase 2, labelled target or non-target by StimulusType. The 16 shared channels were filtered from 0.5 to 30 Hz (fourth-order zero-phase Butterworth), epoched from 200 ms before to 800 ms after each event, baseline corrected to the prestimulus interval, and rejected when absolute amplitude exceeded 150 microvolts; a session entered the analysis only if at least 10 target and 40 non-target epochs survived. Each epoch was then decimated by taking every fourth sample of the 256 Hz signal (64 samples per channel, 1,024 features, 64 Hz effective rate), chosen so the resulting 32 Hz Nyquist frequency stays above the bandpass’s 30 Hz upper edge without folding the retained band; extraction rejects any file sampled below 240 Hz, the rate at which this factor would begin to fold the passband.

The primary score, calibration-derived decoder discriminability, was the out-of-fold area under the receiver operating characteristic curve of an L2-regularised logistic classifier (C = 1.0, lbfgs solver) trained on standardised features, with stratified grouped cross-validation by European Data Format file so no epoch was scored by a model fitted on its own recording file (folds: the smaller of five and the number of files in the session). The score is a property of a session, constant across its recorded conditions, and measures how well that session’s calibration data support a decoder, not an independent physiological marker of user aptitude. This classifier and the linear discriminant comparator are the families evaluated for this paradigm in the literature;[20,21] spatially filtered/Riemannian pipelines[22,23,24] and the wider decoding classifier set[25] were not used, since the study evaluates a calibration-to-accuracy mapping rather than proposing a decoder.

Comparator scores from the same calibration epochs were regularised linear discriminant AUC, grouped cross-validated classification accuracy, mean target-minus-non-target amplitude at Pz (250-500 ms), the same contrast over six posterior channels, and maximum posterior signed r-squared, each run through the identical withheld-cohort procedure (supplement). An exploratory specification adding ALSFRS-R to the primary score is restricted to the 3 of 18 cohorts carrying an observed value, reported alongside the primary score on those records.

### Outcome

At each Test-phase transition to phase 3, the intended character was the final nonzero CurrentTarget in the preceding phase-2 interval and the selected character the modal nonzero SelectedTarget during phase 3. A feedback phase was eligible when feedback was displayed, both values were recoverable, and artificial feedback did not override the selection; correctness required an exact character match. The outcome was the proportion of correct eligible selections within a participant-session-condition record.

### Model Specification

The development model was a logistic regression of character-level correctness on the standardised calibration score, fitted on character-expanded records from the development studies. For a session with score s, estimated accuracy is the inverse logit of a + b (s - m) / d, where m, d are the development mean/SD and a, b the fitted coefficients (fold values in supplement Table S12, so any estimate can be recomputed). A session contributes to the fit in proportion to its characters, so the quantity estimated is the accuracy of a randomly chosen character selection; refitting with sessions, and separately participants, weighted equally moved the pooled error by less than 0.003 (supplement, S3).

### Validation Design

The primary evaluation withheld one source study at a time. Three further analyses used the wider cohort set: an ALS subgroup analysis within the four ALS cohorts alone, a transfer analysis developing the mapping on the other cohorts with every ALS cohort withheld simultaneously, and a moderation analysis testing whether the cohort-specific calibration slope (from a binomial regression of observed correctness fitted within each withheld cohort, clustered on participant) differed between ALS and other cohorts, with the cohort as the unit of comparison since cohort type does not vary within a cohort’s records. Because four cohorts cannot support a single decisive test, it is reported three ways: an unweighted Welch’s t-test on the 18 cohort-specific slopes (primary); an exact permutation test enumerating all 3,060 ALS-label assignments, studentised with the same statistic; and a random-effects meta-regression with Knapp-Hartung inference. The comparison was refitted with each cohort dropped in turn.

### Statistical Analysis

The primary metric was the mean absolute difference between estimated and observed accuracy in the withheld cohort, referenced against a development-mean benchmark, a held-out-cohort-mean benchmark, and skill against the development-mean benchmark (one minus the ratio of their errors; reported against this benchmark throughout unless the cohort’s own mean is named). The cohort’s-own-mean benchmark is harder pooled but easier in 7 of 18 cohorts, since mean absolute error is minimised by the median rather than the mean. Secondary metrics were root mean squared error, character-weighted Brier score, Brier skill score, calibration intercept and slope, and character-weighted AUC, with intercept and slope reported per cohort as well as pooled.[26,27]

Two uncertainty statements are reported and are not interchangeable. Bootstrap confidence intervals (2,000 deterministic replicates, resampling development and withheld participant clusters within source study and refitting) describe uncertainty conditional on the observed cohorts. Separately, treating the source study as the unit of replication, withheld-cohort estimates were summarised by their mean and between-study SD, with a t-distributed interval for the mean and a prediction interval for a cohort not represented in the archive, the quantity a reader should use for their own cohort.[16,17] Cohort-level mean absolute error was pooled on the log scale, since it cannot be negative; the reported mean is the back-transformed geometric mean and its SD stays on the log scale.

For the calibration intercept and slope, a random-effects meta-analysis of the 18 cohort estimates replaced that summary, since cohorts differ in size and the uncorrected SD counts sampling error as between-cohort variation. Tau was estimated by the method of moments[28] with a Q-profile interval,[29] the Paule and Mandel estimate[30] reported alongside it, and the Q test, I^2^[31] and a 95% prediction interval accompany each; all 18 cohorts were fitted and none dropped. Under the participant-cluster bootstrap (Discussion), Study S1’s resamples fell onto the separation boundary often enough that fewer than half of its 2,000 replicates were identified. Repeating cluster-robust pooling on the identical 17 cohorts the bootstrap identified gave tau = 0.44 (slope) and 0.87 (intercept), no closer to the bootstrap’s own 0.37/0.77 than the all-18 values of 0.43/0.87 were, so most of the gap reflects the variance estimator rather than Study S1’s exclusion (supplement, S4).

The primary specification clusters standard errors on participant, since the 739 records come from 271 participants and conditions within one session share a predictor value by construction; treating each record as independent would understate within-cohort variance and inflate the estimated between-cohort variance. A quasi-binomial specification and the uncorrected model-based specification are reported as sensitivities; results agree at the pooled level, though cohort by cohort standard errors differ by up to 40%, with clustering shrinking the standard error in 3 of 18 cohorts.

Each cohort contains between 5 and 24 participants, and a cluster-robust variance is downward-biased below roughly 30 clusters even with the finite-sample correction applied here, which inflates tau and I^2^ in the direction that favours the conclusion: a 20% understatement of the within-cohort variances would put the slope I^2^ below the conventional 75% threshold. I^2^ is therefore reported with its confidence interval as a description only, and no conclusion in this paper rests on its position relative to that threshold; the conclusion is instead checked by refitting at each confidence limit of tau and by repooling with every within-cohort variance inflated by a common factor.

Protocol descriptors were tested as moderators of the cohort calibration slope in the same framework, standardised per between-cohort SD, with Knapp-Hartung inference. Ten were available. The median inter-selection interval, the difference between consecutive selections within a recording file (never across a file boundary), was summarised as the median of per-file medians over all reconstructed selections; a cohort was classed fixed-interval when the relative spread within a file was at most 1e-6, separating a spread of at most 8e-16 from one of at least 0.050. The largest target index and number of distinct targets were empirical proxies for speller-matrix size; grid size and stimulus paradigm were transcribed from the archive’s own data descriptor, with a study coded as checkerboard-paradigm if any documented paradigm was a checkerboard variant (CB, CBcol, sCB): only 2 of 18 studies (Study D, Study J) lacked one. The equipment code (gUSBAmp) is a single archive-wide value, not per-study, so could not be a moderator. P values were corrected across the ten by the Holm procedure. The variance share a moderator accounts for is one minus the ratio of residual tau squared with it to that without, reported as zero where that ratio exceeds one.

Because records are session-conditions nested within sessions within participants, the intraclass correlation of session accuracy within participant is reported alongside an effective number of independent sessions, with associations reported as Pearson and Spearman coefficients at the session, study-centred and participant levels. Precision for the calibration slope is the departure from unity a Wald z test could have detected with 80% power at a two-sided 5% threshold (2.80 times the pooled slope’s standard error); this concerns the average slope, not its between-cohort spread (tau). Four sensitivity analyses, listed before any was run on the widened cohort, covered restriction to cohorts with meaningful outcome variance, exclusion of high-artifact-rejection records, exclusion of small-denominator records, and leave-two-studies-out development (16-cohort development; each cohort in 17 of 153 pairs, its 17 estimates averaged before pooling since a pair shares a cohort with 32 others).

The significance threshold was P < .05, two-sided. Analyses used Python 3.11 with NumPy 2.3, SciPy 1.16, scikit-learn 1.7, statsmodels 0.14, and pandas 2.3.

### Ethics

This was a secondary analysis of BigP3BCI version 1.0.0 (doi:10.13026/0byy-ry86), a publicly available, fully de-identified archive, involving no new data collection, participant contact, or intervention, so institutional review board approval was not required. The original source studies’ ethics approvals and consent statements are reported in the archive documentation.

### What the Calibration Score Is, and What It Is Not

In these copy-spelling protocols the Train phase supplies the data from which the online classifier is derived before the Test phase begins, and the source-study publications that report this practice describe a linear classifier fitted on each session’s calibration data.[32,33] The archive itself names no online classifier for any source study, and the BCI2000 parameter files that would carry its specification were withheld from the release, so the deployed decoder cannot be reconstructed here. The predictor evaluated in this study is in any case not that decoder: it is the out-of-fold discriminability of a separate classifier fitted here on the same calibration recordings. The score is therefore reported as the cross-validated learnability of that session’s calibration data, not as a property of the deployed decoder and not as a physiological marker of user aptitude.

## Results

### Cohort

All 20 documented source studies supplied the shared 16-channel montage at 256 Hz and were screened; 18 contributed at least one eligible online outcome, and two contributed none (Table 1). The analytic set contained 271 study-scoped participants, 410 participant-sessions, 739 participant-session-condition records, and 19,611 online character selections, of the 19,688 eligible selections in Table 1; the 77-selection difference is reconciled cohort by cohort in Table S1. The four ALS cohorts contributed 47 participants, 113 sessions, 194 records, and 3,318 selections. Observed session-condition accuracy averaged 0.851, and 228 of 739 records (30.9%) were at 100%, with three cohorts near ceiling (0.963 to 0.997).

Because the calibration score is a property of a session, the 739 records carry 410 distinct predictor values; session accuracy clustered within participant (intraclass correlation 0.412), giving approximately 338 effective independent sessions.

### Association Between Calibration-Derived Decoder Discriminability and Online Accuracy

**The point estimate was positive in all 18 contributing cohorts, but its magnitude varied widely and several cohort-specific estimates were imprecise, with confidence intervals including values close to zero (Table S6); pooled across sessions the association was precisely estimated, at r = 0.677 (95% CI 0.621 to 0.726) once both variables are centred within cohort.** Within-cohort Pearson r ran from 0.190 to 0.928 (median 0.635), and the 95% confidence interval included zero in 6 of the 18 cohorts (widest: -0.42 to 0.86). Pooling sessions gave r = 0.716 (95% CI 0.665 to 0.760, n = 410, p < 0.001); centring both variables within cohort, which removes cohort means but not differences in variance or precision, gave r = 0.677 (0.621 to 0.726, p < 0.001), so the association is not produced by cohorts differing in mean difficulty and signal quality. Collapsing each participant to a single observation gave r = 0.714 (0.650 to 0.768, n = 271, p < 0.001; Spearman □ = 0.756).

### Estimation Error and Its Reference Points

#### Pooled across withheld cohorts, estimated session accuracy fell about a third closer to observed accuracy than the development-mean benchmark, and also beat the benchmark of each cohort’s own mean

Across withheld cohorts the mean absolute error was 0.098 (95% CI 0.091 to 0.107), against 0.146 for the development-mean benchmark and 0.123 for the held-out-cohort-mean benchmark, giving a skill of 0.327. This pools every withheld record and weights larger cohorts more heavily; the mean of the 18 cohort-level errors, reported next, is a different quantity. The character-weighted Brier score was 0.123 (95% CI 0.113 to 0.132), Brier skill 0.110 (95% CI 0.063 to 0.149), and character-weighted AUC 0.748 (95% CI 0.719 to 0.770); these intervals are conditional on the 18 observed cohorts.

### Transportability

#### Treating the source study as the unit of replication, discrimination transported on average but not dependably, and calibration did not transport (Figures 1 and 2)

Across the 18 withheld cohorts the mean absolute error, pooled on the log scale, averaged 0.090 (geometric mean; arithmetic mean 0.101) with a between-cohort SD of 0.477 on the log scale, giving a 95% interval for the mean of 0.071 to 0.115 and a new-cohort interval of 0.032 to 0.254 (Figure 3). AUC averaged 0.713 across cohorts (new-cohort interval 0.491 to 0.935, lower bound chance); Brier skill averaged 0.174 (new-cohort interval -0.296 to 0.644). Every study-level summary here is tabulated in Table S3.

**Figure 1.**
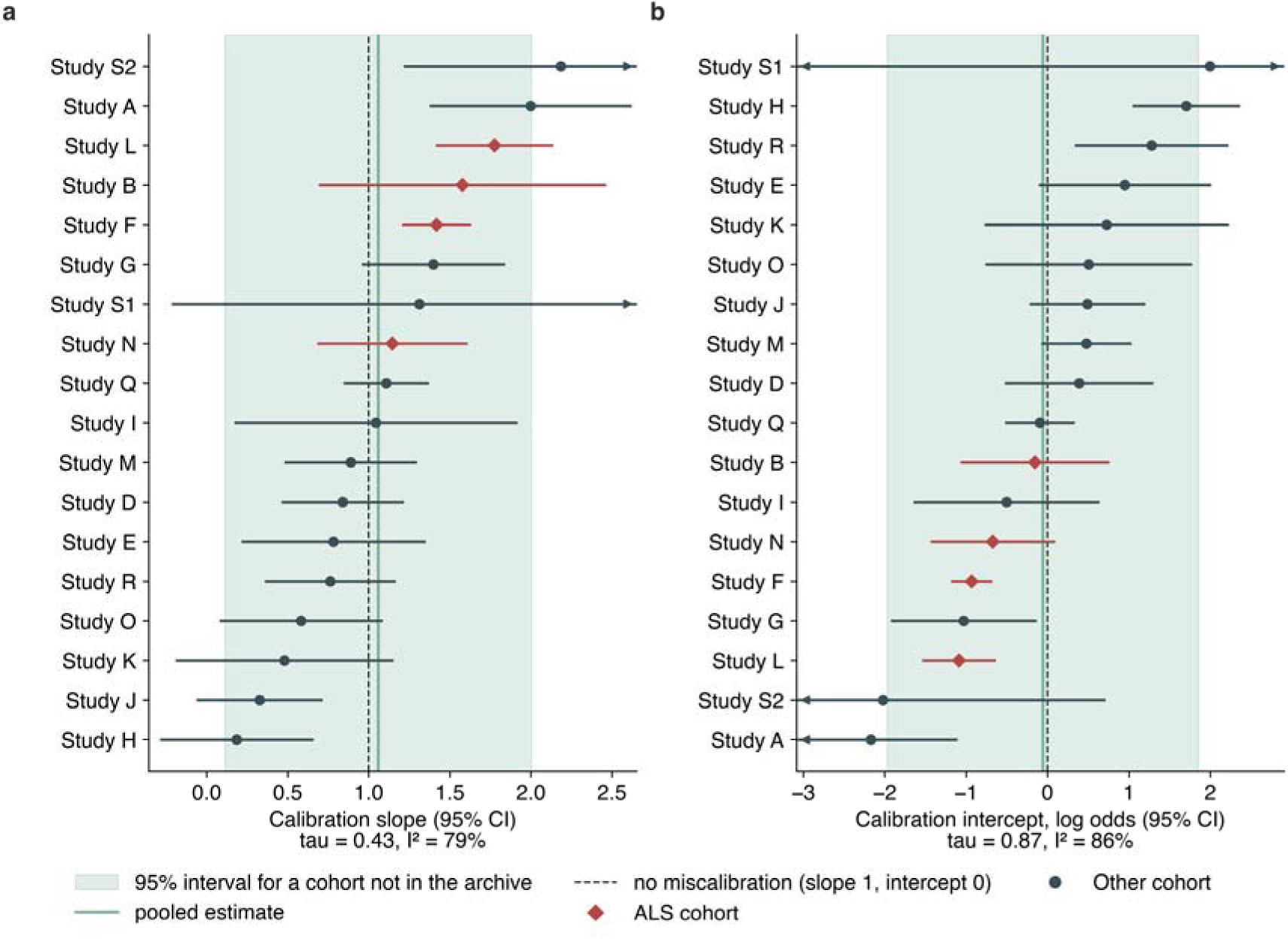
Calibration slope and intercept in each withheld cohort, against the values a transportable mapping would take. Each row is one cohort withheld from model development, with its estimate and 95% confidence interval, marked by whether the archive documents an ALS study population; standard errors are clustered on participant. Each panel is ordered by its own estimate. The dashed line is the value a mapping that transported exactly would take (one for slope, zero for intercept); the band is the 95% interval for a cohort not represented in the archive, computed at tau’s point estimate, and the solid line within it is the pooled estimate. Intervals running past the axis are drawn with an arrow. Tau and I^2^ are given beneath each panel; I^2^ is descriptive only (Methods, Statistical Analysis).

**Figure 2.**
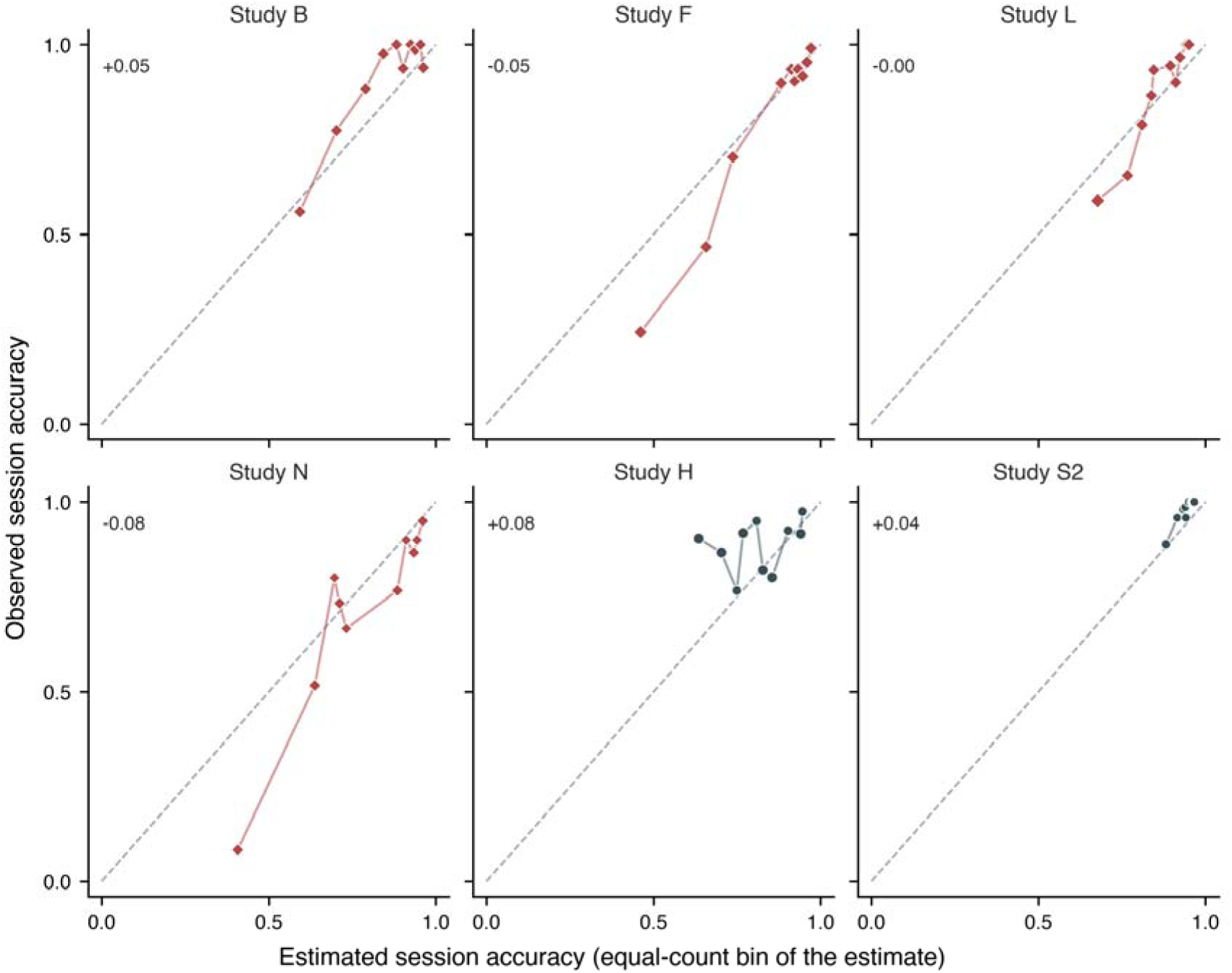
Observed against estimated session accuracy, in the four ALS cohorts and the two non-ALS cohorts at either end of the calibration-slope range (Figure 1). Each point is one of up to ten equal-count bins of the estimate within that cohort, placed at the selection-weighted mean estimate and observed accuracy, with point area increasing with the selections it rests on. The dashed line is equality; points above it are bins the mapping understated, points below are bins it overstated. The number in each panel is observed minus estimated accuracy across that cohort. The full 18-cohort version is Figure S2.

**Figure 3.**
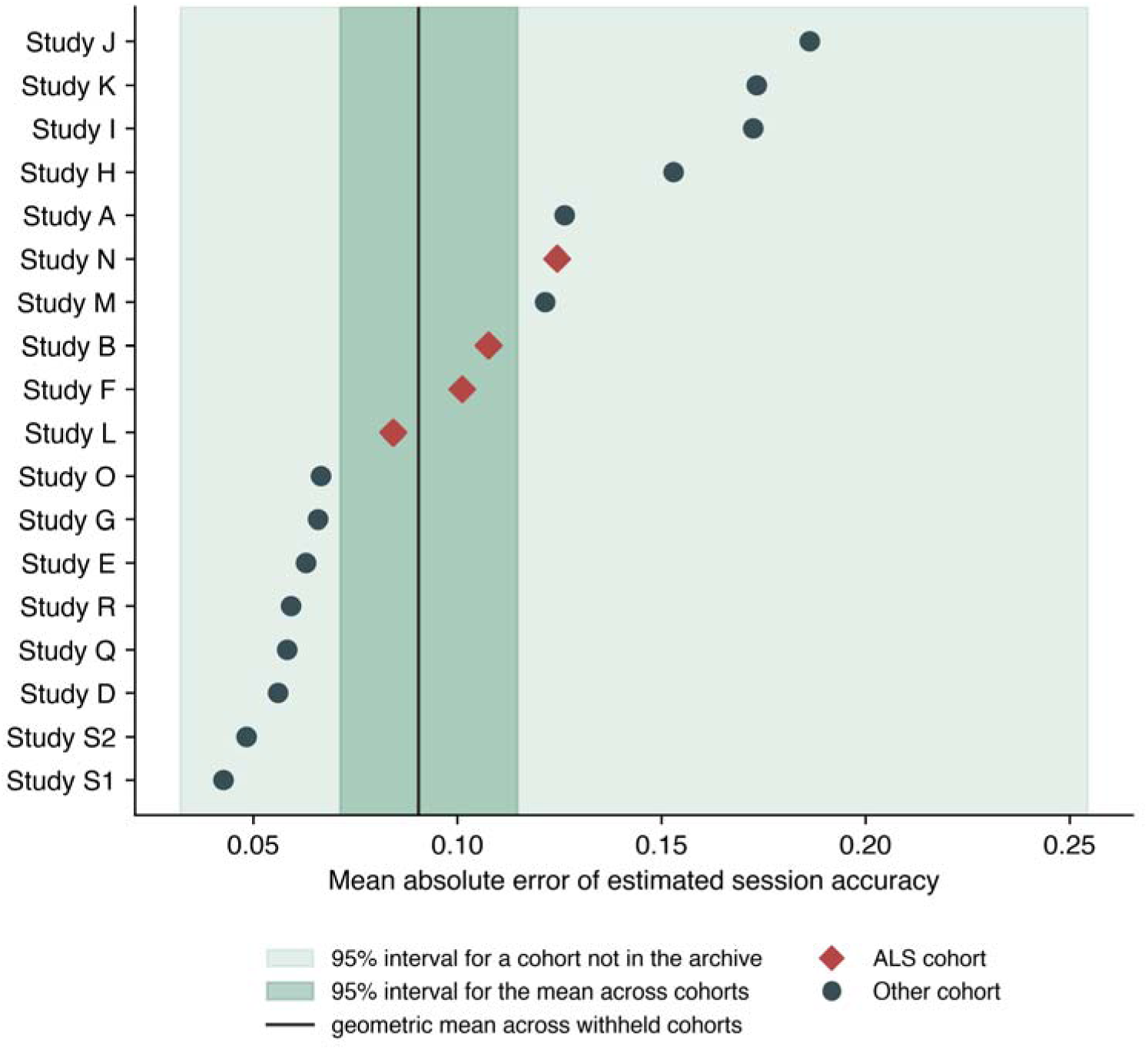
Estimation error in each withheld cohort, with the two uncertainty statements. Each point is one cohort withheld from model development. The darker band is the 95% interval for the geometric mean across observed cohorts; the lighter band is the 95% interval for a cohort not represented in the archive. Only the second describes what a reader should expect in their own setting.

#### The calibration intercept varied so widely between cohorts that a single fitted mapping was displaced in many of them

It departed from zero by at least half a log-odds unit in 13 of the 18 cohorts and by at least a full unit in 7, with its 95% interval excluding zero in 6. Under the primary cluster-robust specification the between-cohort SD of the calibration intercept was tau = 0.87 (95% CI 0.60 to 1.51) against a pooled intercept of -0.06 (Q = 122.6, df = 17, p < 0.001); I^2^ was 86.1 (95% CI 79.5 to 90.6), a description only (Methods, Statistical Analysis). The observed intercepts ran from -2.169 to 1.995, and the 95% interval for a cohort not represented in the archive runs from -1.97 to 1.85 on the log-odds scale (Figure 1), the difference between a mapping that badly understates and one that badly overstates accuracy. This holds regardless of where in its interval tau lies: at tau’s lower confidence limit the new-cohort interval is -1.39 to 1.26, and it is -1.37 to 1.06 with every within-cohort variance inflated fivefold.

#### The calibration slope varied in the same direction, and its between-cohort variance is itself imprecisely estimated

tau = 0.43 (95% CI 0.30 to 0.77) against a pooled slope of 1.06 (95% CI 0.82 to 1.30), Q = 81.3 (df = 17, p < 0.001), I^2^ 79.1 (95% CI 67.6 to 86.5), an interval that spans the same threshold, so no claim rests on it either. The observed slopes ran from 0.185 to 2.185 and the new-cohort interval runs from 0.111 to 2.005 (Figure 1); at tau’s lower confidence limit it is 0.394 to 1.704, across which no single fitted mapping is usable, and it excludes zero at tau’s point estimate but includes it at tau’s upper limit (-0.603 to 2.759), so that exclusion is a property of one value of tau rather than a finding. Cohort by cohort, 4 of the 18 slope confidence intervals include zero (Figure 1). Fitting one calibration curve to all withheld predictions pooled gives a slope of 0.967 (95% CI 0.791 to 1.127) and an intercept of 0.054 (95% CI -0.264 to 0.395), averaging opposing departures visible separately in Figure 2.

**Table 2.** Per-cohort composition and observed performance. For each of the 18 contributing cohorts: participants, records, analysed selections, mean observed accuracy, and, when withheld from development, mean absolute error and character-weighted AUC. ALS cohorts are listed first. Calibration intercept and slope with their confidence intervals are shown in Figure 1 rather than tabulated here; the full per-cohort table, also carrying grid size and checkerboard-paradigm indicators, is Table S2.

| Cohort | Participants | Records | Selections | Observed accuracy | MAE | AUC |
| --- | --- | --- | --- | --- | --- | --- |
| Study B | 18 | 56 | 781 | 0.899 | 0.108 | 0.825 |
| Study F | 10 | 89 | 1,067 | 0.778 | 0.101 | 0.863 |
| Study L | 11 | 33 | 990 | 0.839 | 0.084 | 0.811 |
| Study N | 8 | 16 | 480 | 0.694 | 0.124 | 0.814 |
| Study A | 13 | 39 | 1,404 | 0.786 | 0.126 | 0.775 |
| Study D | 17 | 34 | 1,230 | 0.896 | 0.056 | 0.641 |
| Study E | 8 | 8 | 240 | 0.921 | 0.063 | 0.668 |
| Study G | 20 | 40 | 1,198 | 0.886 | 0.066 | 0.732 |
| Study H | 16 | 64 | 1,926 | 0.877 | 0.153 | 0.520 |
| Study I | 13 | 26 | 948 | 0.658 | 0.172 | 0.674 |
| Study J | 20 | 40 | 1,812 | 0.708 | 0.186 | 0.600 |
| Study K | 5 | 16 | 480 | 0.750 | 0.173 | 0.639 |
| Study M | 21 | 42 | 1,260 | 0.823 | 0.121 | 0.685 |
| Study O | 17 | 34 | 1,187 | 0.884 | 0.067 | 0.581 |
| Study Q | 20 | 54 | 1,944 | 0.820 | 0.058 | 0.682 |
| Study R | 20 | 80 | 1,440 | 0.963 | 0.059 | 0.650 |
| Study S1 | 10 | 20 | 360 | 0.997 | 0.043 | 0.851 |
| Study S2 | 24 | 48 | 864 | 0.976 | 0.048 | 0.826 |

A small average departure of the slope from unity was not within reach of this design: the pooled slope’s standard error of 0.122 means a Wald z test against unity would have detected a departure of at least 0.34 with 80% power, and its confidence interval has a half-width of 0.24. Neither bears on transportability, since what a single fitted mapping has to survive is the slope’s spread across cohorts, and the design placed tau between 0.30 and 0.77 without resolving where in that range it lies.

The uncorrected standard deviations of the 18 cohort estimates (supplement, Table S3) are 0.560 for the slope and 1.175 for the intercept; both exceed tau because they count sampling error as between-cohort variation, and tau is used in preference throughout.

#### One cohort was estimated less accurately than the development-mean benchmark, and six were estimated less accurately than their own cohort mean

Skill against the development-mean benchmark was negative in one of 18 cohorts (Figure 4). Against the harder benchmark of each cohort’s own mean, skill was negative in six of 18 (Table S8): one cohort underperformed both benchmarks, four had observed accuracy so high (0.921 to 0.997) that their own mean already tracked nearly every record, and one showed a negligible (-0.008) shortfall rather than a real one. All four ALS cohorts had positive skill against both benchmarks, at 0.317, 0.383, 0.448 and 0.502 against the development mean.

**Figure 4.**
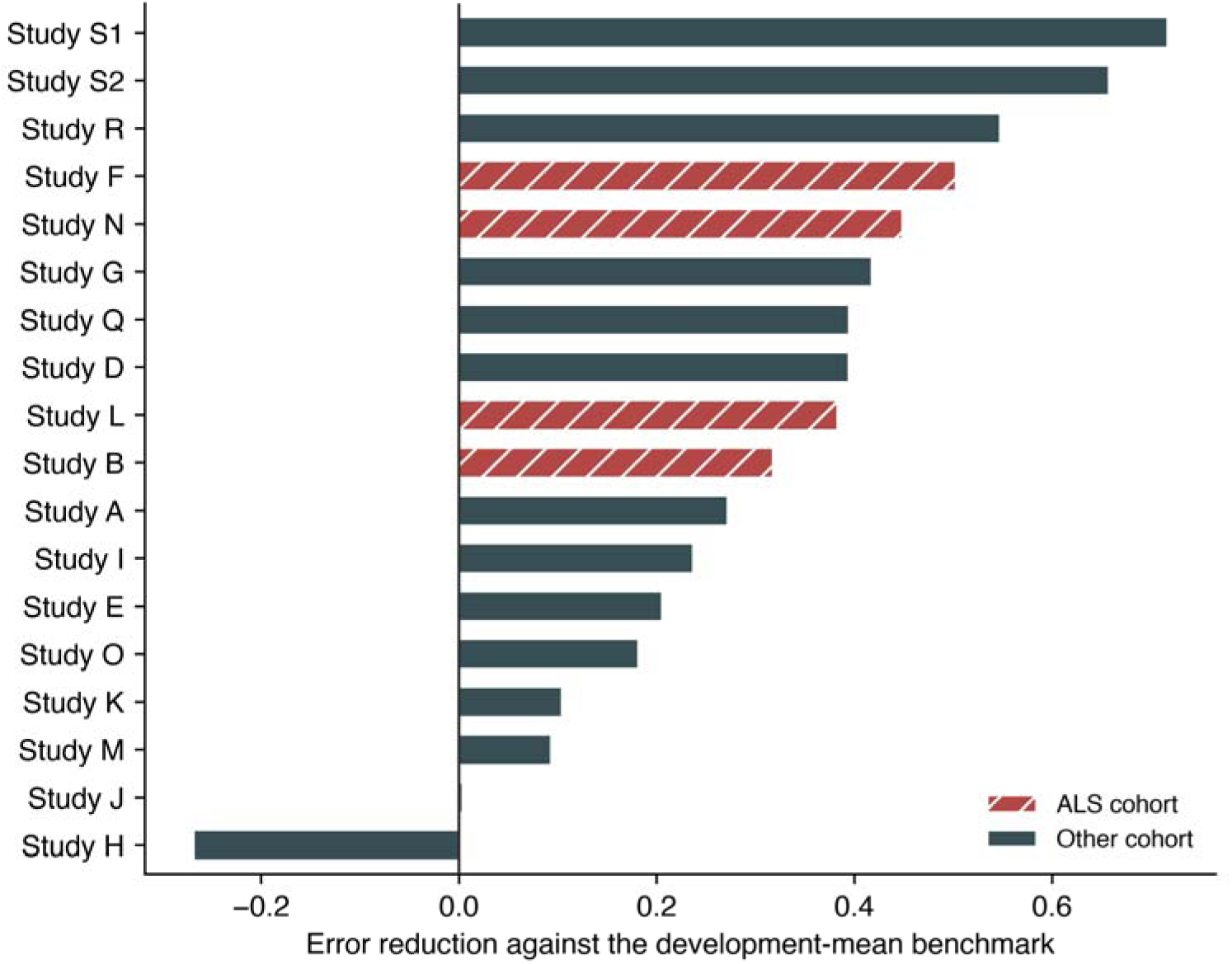
Error reduction in each withheld cohort against the development-mean benchmark. Values below zero indicate the calibration score estimated accuracy less well than the development-set mean. All four ALS cohorts were positive; one non-ALS cohort was negative.

### Amyotrophic Lateral Sclerosis Subgroup

#### Restricting the analysis to the four ALS cohorts reproduced a more favourable and considerably more precise picture than the full archive supported

Within this subgroup the mean absolute error was 0.091 (95% CI 0.076 to 0.108), Brier skill 0.303 (95% CI 0.184 to 0.424), AUC 0.831 (95% CI 0.768 to 0.869), calibration intercept 0.025 (95% CI -0.240 to 0.341), and slope 0.985 (95% CI 0.786 to 1.188). The uncorrected between-cohort SD of the calibration slope was 0.223 within this subgroup against 0.560 across all cohorts; four cohorts are too few to estimate tau usefully, so the uncorrected spread is used on both sides.

### Transfer From Cohorts Without a Documented ALS Population

#### A mapping developed without any ALS data estimated accuracy in the ALS cohorts with modest loss

With all four ALS cohorts withheld from development simultaneously, mean absolute error in each was 0.087, 0.106, 0.109, and 0.131 (mean 0.108), against 0.104 when other ALS cohorts were available; signed bias ranged from -0.086 to 0.049.

### Cohort Type as a Moderator

#### With the cohort as the unit of analysis, calibration slope was higher in the ALS cohorts, but not precisely enough to support a claim

The cohort-specific slope averaged 1.479 in the four ALS cohorts (SD 0.266) and 0.992 in the other 14 (SD 0.580), a difference of 0.487 (SE 0.204; 95% CI 0.040 to 0.933; Welch t = 2.38, df = 11.7, p = 0.035). An exact permutation test gave p = 0.058; a random-effects meta-regression gave a difference of 0.559 (95% CI -0.009 to 1.127, p = 0.053), leaving a residual between-cohort SD of 0.349 against 0.432 with cohort type removed, so cohort type accounts for about a third of the between-cohort variance and most remains unexplained.

### The comparison also turns on individual cohorts

Dropping one ALS cohort moves the Welch p value to between 0.008 and 0.111, and dropping one of the other 14 moves it to between 0.013 and 0.060; all three tests agree in direction and magnitude and fall on either side of the conventional threshold. These are the leave-one-cohort-out slopes, not the pooled 0.985 reported for the ALS subgroup above (developed within the ALS cohorts alone); for the same reason their SD of 0.266 is not the 0.223 reported there, since 0.266 is the ALS spread under the primary design, comparable with the 0.580 of the other 14.

Cohort type is a study-level attribute entangled with paradigm, hardware and stopping rule; only four studies carry a documented ALS population and the rest are not documented as healthy controls, so this comparison is exploratory and cannot separate population from protocol (records shown by cohort type in Figure S1, without a fitted line, Methods).

### Protocol Descriptors as Moderators

#### The time a cohort took over each selection was associated with a two-thirds reduction in estimated residual between-cohort variance in calibration slope

Ten descriptors were entered one at a time as moderators in a random-effects meta-regression of the cohort slope, weighted by precision, standardised per between-cohort SD, with Holm-corrected p values; all 18 cohorts were fitted for every descriptor. The strongest was the median inter-selection interval (7.3 to 50.0 seconds across cohorts, the archive’s only trace of the stopping rule): 0.406 per SD (95% CI 0.181 to 0.632; t = 3.82, df = 16, p = 0.002, Holm p = 0.015), reducing the between-cohort variance of the slope from tau squared = 0.187 to 0.063, a share of 66% (residual SD 0.25 against 0.43). The rank correlation agreed at rho = 0.72 (p < 0.001, Holm p = 0.007), and on the log scale the share was 71% (p < 0.001); across 18 leave-one-cohort-out refits its uncorrected p value never exceeded 0.010 and its share ran from 54% to 82%. Cohorts that spent longer per selection had steeper slopes; the reported effect of stimulus repetition is on accuracy rather than on the slope,[32,34,35] and the interval does not track cohort mean accuracy here (rho = 0.16, p = 0.52), so the direction is not read from that work. A mechanism is available (accumulated evidence grows with the square root of repetitions), but is not tested here.

No other descriptor was associated with a share of the between-cohort variance distinguishable from zero, including the archive’s documented grid size, checkerboard-variant paradigm, and both empirical matrix-size proxies (supplement, S6); the result was also robust to how the stopping-rule descriptor was defined, whether it partly reflects the recording rather than a fixed protocol setting, and how the multiple-comparison correction was applied (supplement, S6).

### Predictor From a Preceding Session

#### Using a calibration recording from a preceding session, rather than from the session being estimated, attenuated but did not remove the association

Among 139 consecutive session pairs, the earlier session’s score correlated with the later session’s accuracy at r = 0.513 (95% CI 0.379 to 0.626, p < 0.001), against r = 0.728 (0.639 to 0.798) for the score recorded in the session being estimated.

### Sensitivity Analyses

Results were consistent under the four checks named in the Methods: excluding low-count records, restricting to cohorts with meaningful accuracy variation, excluding heavy-calibration-rejection records, and leave-two-studies-out development (16 rather than 17 cohorts, reproducing the primary result across all 153 splits; Table S7). The cohort-subset comparison restricted to cohorts without a documented ALS population, shown alongside these checks in Table S7, also gave a consistent result. Every comparator predictor named in the Methods is reported in full in Table S9; the regularised linear discriminant agreed with the primary score to within 0.007 on most columns, the two exceptions being the per-cohort slope range and the between-cohort MAE SD, so the transportability result reflects how separable the calibration data are rather than the classifier family used to measure it.

## Discussion

Across 18 source-study cohorts, the discriminability of a classifier fitted to a session’s calibration block was related to that same session’s online spelling accuracy, with a positive point estimate in every cohort, at the participant level, and when the calibration recording came from an earlier session. A fitted mapping from that score to expected accuracy did not transport: the calibration intercept had tau = 0.87 (95% CI 0.60 to 1.51) and an unrepresented-cohort interval of -1.97 to 1.85 log-odds; slope heterogeneity gave the same conclusion, at tau = 0.43 (0.30 to 0.77) and a range of 0.185 to 2.185. One cohort was estimated less accurately than the development-mean benchmark, six less accurately than their own mean.

That distinction determines what a calibration score can be used for. Ranking sessions within a setting, which requires only that the association hold locally, is supported. Reporting expected accuracy in a cohort where the mapping was not developed is not supported without local recalibration, since the interval for a cohort outside this archive spans 0.032 to 0.254 for estimation error and includes chance-level discrimination and negative skill.

Mainsah and colleagues derived speller accuracy analytically from a calibration-derived detectability index and validated it within study;[15] this analysis is the transportability counterpart to that work, reproducing the association with a positive point estimate in every cohort and adding that the numerical mapping is cohort-specific. Earlier reports relating calibration measures to P300 performance in ALS[11,12] and to brain-computer interface performance generally[10] are likewise consistent with the association reported here.

Predictor precision differs between cohorts (standard error 0.008 to 0.031, a fourfold spread), and measurement error attenuates a fitted slope, so some between-cohort spread could reflect precision. Correcting each cohort’s slope for its reliability (0.848 to 0.992) moved tau only from 0.432 to 0.422, surviving a stress test on the assumed measurement-error variance (supplement, S9): measurement error is real but minor, and does not explain the mapping’s failure to transport.

Part of the variation is attributable to protocol rather than to the cohorts themselves: the time each cohort took over a selection, the archive’s only trace of the stopping rule, was associated with the two-thirds reduction in residual between-cohort variance reported above (Results, Protocol Descriptors as Moderators). That association does not restore transportability, since a mapping whose slope depends on the stopping rule still cannot be carried to a site whose stopping rule is unknown, but it means the residual spread is not an irreducible population property, and a site reporting its stopping rule alongside a calibration score would remove a substantial share of the uncertainty in a transported estimate.

The 18 development folds are themselves stable: their intercept and slope coefficients (supplement, Table S12) vary by a coefficient of variation of 2.5% and 3.3% across folds, against 49.4% for the held-out cohort-specific slopes in Table S2, evidence against development-fit noise as an explanation for the heterogeneity (the between-cohort spread reflects how each fold performs in its withheld cohort, not instability in what each fold fits).

The 18 folds are also not independent, since any two share up to 16 of 17 development cohorts, and a joint bootstrap addresses that correlation directly: resampling every cohort’s participants once per replicate and refitting all 18 folds from that single draw gave a within-replicate between-cohort SD of the slope averaging 0.79 across 2,000 replicates (95% range 0.49 to 1.56) and of the intercept 1.62 (0.94 to 3.36), larger than the meta-analytic tau (0.43, 0.87). This is expected rather than discordant: since the procedure never resamples which 18 cohorts are observed, only participants within each fixed cohort, it mixes genuine heterogeneity with within-cohort sampling noise and estimates something closer to the square root of tau squared plus mean within-cohort sampling variance, structurally at least as large as tau, not a corrected or dependence-aware version of tau itself (supplement, S4).

The four ALS cohorts alone gave a more favourable and homogeneous picture than the full archive (Results, ALS Subgroup); evaluating a mapping on few withheld cohorts should be expected to understate how much performance varies elsewhere.

One analysis improved transportability: excluding records with more than 20% of calibration epochs above the artifact threshold reduced estimation error from 0.090 to 0.081 and the uncorrected between-cohort SD of the calibration slope from 0.560 to 0.465. Screening calibration data quality is therefore a candidate step warranting prospective evaluation, available at no cost since the rejection fraction is computed alongside the score.

A mapping developed entirely without ALS data estimated ALS-cohort accuracy with a mean absolute error of 0.108, against 0.104 with other ALS cohorts available, so the relationship is not absent outside the clinical population, though whether it is population-dependent in another sense is unresolved. The calibration slope averaged higher in the ALS cohorts, one possible mechanism for mis-calibration, but with the cohort as the unit of analysis that difference is imprecise, the three tests fall on either side of the conventional threshold, a single cohort moves it, and cohort type cannot be separated from the paradigm, hardware and stopping rule that differ alongside it.

### Study Limitations

First, calibration and online blocks come from the same session throughout; temporal precedence cannot be verified from de-identified timestamps, so the separation enforced is between protocol phases, not demonstrated ordering (the preceding-session analysis is the closest approximation and shows an attenuated association).

Second, the predictor is not the deployed decoder (Methods, What the Calibration Score Is, and What It Is Not).

Third, the cohorts differ in speller matrix, stimulus paradigm, electrode type, and stopping rule, and paradigm is largely nested within source study, so withholding a cohort also withholds its paradigms and the two effects cannot be separated. Two protocol traces were tested (Results, Protocol Descriptors as Moderators): the time taken per selection, and the range of target indices; the archive’s documented grid size and stimulus paradigm were tested directly too, and none was associated with a share distinguishable from zero. What remains untestable is the stopping algorithm’s thresholds, the exact grid layout beyond documented size, and the online decision procedure.

Fourth, 30.9% of records were at 100% accuracy and three cohorts were near ceiling; the sensitivity analysis restricted to cohorts with meaningful outcome variance gave a higher estimation error, so the primary estimate is favourably influenced by cohorts where the task was easy.

Fifth, the outcome is character-level selection accuracy, an operational endpoint rather than communication effectiveness, quality of life, or any clinical outcome, and does not capture communication rate.[36] Sixth, these are legacy protocols; the analysis does not establish performance with contemporary assistive-communication workflows. Seventh, the archive identifies an ALS population for four cohorts only, with no participant-level clinical characteristics available for any cohort.

Eighth, the cohort-type comparison carries two further limitations: each cohort’s slope comes from a model developed on the other 17 (14 without a documented ALS population), so part of the higher ALS slope could reflect development-set composition rather than population, and the 18 slopes share most of their development data, correlating them positively and making all three tests’ standard errors somewhat optimistic. Neither can be addressed at this number of cohorts.

Ninth, the 18 held-out estimates are not fully independent, each development fold sharing 16 of the other 17 cohorts; the joint bootstrap above measures rather than merely bounds this correlation, and the fold-coefficient stability reported there is evidence against it inflating the observed spread.

Tenth, the between-cohort variance is itself estimated with limited precision (18 cohorts of 5 to 24 participants), and a cluster-robust variance at these cluster counts is downward-biased, inflating tau and I^2^ in the direction that favours the conclusion (Methods, Statistical Analysis); the conclusion holds at tau’s lower confidence limit and under the participant-cluster bootstrap, which gave a smaller tau of 0.37 (slope) and 0.77 (intercept) (supplement, S4).

Eleventh, because participant identifiers are study-scoped, we could not determine whether any individuals participated in more than one source study.

## Conclusion

In 18 source-study cohorts, calibration-derived decoder discriminability was related to online spelling accuracy in the corresponding session, with a positive point estimate in every cohort, but the fitted mapping did not transport: the calibration intercept spanned mappings that badly understate and badly overstate accuracy, the slope varied more than tenfold, and the prediction interval for a cohort outside the archive does not exclude chance discrimination or negative skill. The score may be useful for within-setting ranking after local validation, and the artifact-rejection fraction is a candidate data-quality screen warranting prospective evaluation; neither use has been prospectively validated here, and the score should not be used to report expected accuracy in a cohort where the mapping was not developed without local recalibration. Prospective evaluation would need to predefine the recalibration procedure and measure user-centred communication outcomes rather than character accuracy alone.

## Data Availability

All data produced are available online at https://github.com/Alon-Gorenshtein/P300-speller-performance-in-amyotrophic-lateral-sclerosis-In-BigP3BCI-

## Ethics approval

Institutional review board approval was not required; the determination is stated in full in the Methods, under Ethics.

## Author contributions

Contributions are described using the CRediT taxonomy. A.G.: conceptualization, methodology, software, formal analysis, data curation, validation, visualization, and writing of the original draft. Y.A.: software, data curation, formal analysis, validation, and review and editing of the manuscript. T.L.: investigation, validation, and review and editing of the manuscript. Y.B.: methodology, validation, and review and editing of the manuscript. E.K.: conceptualization, methodology, supervision, resources, and review and editing of the manuscript. O.D.: conceptualization, methodology, investigation, formal analysis, clinical interpretation, supervision, writing of the original draft, and review and editing of the manuscript. All authors critically reviewed the manuscript and approved the final version submitted for publication. A.G. (corresponding author) had full access to all data in the study and takes responsibility for the integrity of the data and the accuracy of the analysis.

## Data availability statement

BigP3BCI version 1.0.0 is publicly available (doi:10.13026/0byy-ry86). Analysis code and frozen outputs are available at https://github.com/Alon-Gorenshtein/P300-speller-performance-in-amyotrophic-lateral-sclerosis-In-BigP3BCI-; a versioned archive with its own DOI will be minted via Zenodo upon acceptance and this citation updated accordingly. The archive is not redistributed.

