## appendix for "Calibration-derived decoder discriminability is associated with online P300-speller accuracy, but the fitted mapping does not transport across cohorts"

### Supplementary material

#### S1. Data provenance

This retrospective secondary analysis used the BigP3BCI version 1.0.0 public archive. The downloaded archive had SHA256 digest eea294aa34e9ed11e5a25d07e30aeefdf8b2d467a8309e2c38405a289afcd72f. Before any signal was processed, the ingestion pipeline checked this archive digest, read the distributor checksum manifest, selected non-AppleDouble European Data Format files, and checked every selected file against its manifest digest. The source archive and cache are not redistributed in this package.

All 20 documented source studies were examined. Every study supplied the 16 shared EEG channels sampled at 256 Hz together with the StimulusBegin, StimulusType and PhaseInSequence event channels, so no study was excluded for montage or event-channel incompatibility. Study-scoped participant identifiers were retained to prevent accidental cross-study linkage; they do not identify unique people across studies.

#### S2. Predictor and outcome reconstruction

The predictor used Train-phase files only. A calibration event was a rising StimulusBegin transition during phase 2, labelled by StimulusType. The 16 shared channels were filtered from 0.5 to 30 Hz with a fourth-order zero-phase Butterworth filter, epoched from 200 ms before to 800 ms after the event, baseline corrected to the prestimulus interval, and rejected at an absolute amplitude above 150 microvolts. A session entered the analysis only when at least 10 target and 40 non-target epochs survived rejection.

Epochs were decimated by taking every fourth sample. At 256 Hz this leaves 64 samples per channel and 1,024 features, and an effective sampling rate of 64 Hz. The resulting Nyquist frequency of 32 Hz stays above the 30 Hz upper edge of the bandpass, so the retained band is not folded onto lower frequencies, and the factor was chosen for that reason. Extraction rejects any file sampled below 240 Hz, the rate at which a factor of four would begin to fold the passband. The bandpass is a fourth-order zero-phase Butterworth rather than a brick wall, so attenuated content above 32 Hz does still fold onto 24 to 32 Hz; the filter response is 8.9 dB down at 32 Hz, that shoulder carries about 1.2% of output power for a white input, and mains at 60 Hz arrives about 60 dB down.

The number of cross-validation folds was the smaller of five and the number of Train files in the session. Fold count therefore differs between sessions and between studies, so the sampling precision of the predictor is not identical across the cohorts whose transportability is being compared.

For each Test-phase transition to phase 3, the intended character was the final nonzero CurrentTarget in the directly preceding phase-2 interval and the selected character was the modal nonzero SelectedTarget during phase 3. A trial was eligible when feedback was displayed, both values could be recovered, and FakeFeedback did not override the selection. Correctness required an exact character match.

**Table S1. Feedback-phase reconstruction by source study.** Eligible counts are at the feedback-phase level. Records also require a usable calibration feature set for the participant-session, which is why 19,611 selections enter the analysis rather than 19,688. The 77-selection difference is two participant-session-condition groups, both in Study B and both from participant B_01: sessions SE002 and SE003 were archived with 10 Test-phase recordings each and no Train-phase recording at all, so those sessions have no calibration epochs and no calibration score, and their 38 and 39 eligible selections cannot be estimated by a model whose predictor is that score. No other cohort loses a selection between the two totals. One further session, in Study C, was excluded at feature extraction because a single calibration file cannot support grouped cross-validation, but Study C contributes no eligible online outcome in any case, so that exclusion does not move either total.

| Source study | Reconstructed | Eligible | Dominant exclusion reason |
| --- | --- | --- | --- |
| Study A | 1,404 | 1,404 | none |
| Study B | 858 | 858 | none |
| Study C | 5,680 | 0 | artificial feedback override |
| Study D | 1,230 | 1,230 | none |
| Study E | 240 | 240 | none |
| Study F | 1,079 | 1,067 | feedback not displayed |
| Study G | 1,198 | 1,198 | none |
| Study H | 1,926 | 1,926 | none |
| Study I | 948 | 948 | none |
| Study J | 1,812 | 1,812 | none |
| Study K | 480 | 480 | none |
| Study L | 990 | 990 | none |
| Study M | 1,260 | 1,260 | none |
| Study N | 480 | 480 | none |
| Study O | 1,202 | 1,187 | feedback not displayed |
| Study P | 2,263 | 0 | intended character not recoverable |
| Study Q | 3,888 | 1,944 | feedback not displayed |
| Study R | 1,440 | 1,440 | none |
| Study S1 | 360 | 360 | none |
| Study S2 | 864 | 864 | none |
| Total | 29,602 | 19,688 |  |

**Table S2. Per-cohort composition, documented protocol metadata, and withheld-cohort performance.** For each of the 18 contributing cohorts: participants, records, analysed selections, the archive's own documented speller grid size and whether it uses a checkerboard-variant stimulus paradigm, mean observed session-condition accuracy, and, when that cohort was withheld from model development, the mean absolute error, character-weighted area under the curve, and calibration intercept and slope, with the slope's 95% confidence interval given as separate lower and upper columns rather than combined into one cell. Calibration intercepts and slopes are from the primary cluster-robust specification, with standard errors clustered on participant. ALS cohorts are listed first. This is the full version of the main-text Table 2, which reports composition and observed performance only; the calibration intercept and slope for each cohort, with their confidence intervals, are also shown in Figure 1.

| Cohort | Participants | Records | Selections | Grid size | Checkerboard | Accuracy | MAE | AUC | Calib. intercept | Calib. slope | Slope CI low | Slope CI high |
| --- | --- | --- | --- | --- | --- | --- | --- | --- | --- | --- | --- | --- |
| Study B | 18 | 56 | 781 | 36 | Yes | 0.899 | 0.108 | 0.825 | -0.154 | 1.578 | 0.691 | 2.464 |
| Study F | 10 | 89 | 1,067 | 72 | Yes | 0.778 | 0.101 | 0.863 | -0.932 | 1.418 | 1.205 | 1.631 |
| Study L | 11 | 33 | 990 | 36 | Yes | 0.839 | 0.084 | 0.811 | -1.088 | 1.775 | 1.413 | 2.137 |
| Study N | 8 | 16 | 480 | 36 | Yes | 0.694 | 0.124 | 0.814 | -0.671 | 1.145 | 0.681 | 1.609 |
| Study A | 13 | 39 | 1,404 | 72 | Yes | 0.786 | 0.126 | 0.775 | -2.169 | 1.999 | 1.375 | 2.622 |
| Study D | 17 | 34 | 1,230 | 72 | No | 0.896 | 0.056 | 0.641 | +0.389 | 0.839 | 0.461 | 1.217 |
| Study E | 8 | 8 | 240 | 72 | Yes | 0.921 | 0.063 | 0.668 | +0.950 | 0.782 | 0.214 | 1.351 |
| Study G | 20 | 40 | 1,198 | 72 | Yes | 0.886 | 0.066 | 0.732 | -1.029 | 1.399 | 0.957 | 1.840 |
| Study H | 16 | 64 | 1,926 | 72 | Yes | 0.877 | 0.153 | 0.520 | +1.705 | 0.185 | -0.289 | 0.659 |
| Study I | 13 | 26 | 948 | 72 | Yes | 0.658 | 0.172 | 0.674 | -0.504 | 1.044 | 0.170 | 1.917 |
| Study J | 20 | 40 | 1,812 | 36 | No | 0.708 | 0.186 | 0.600 | +0.491 | 0.326 | -0.063 | 0.716 |
| Study K | 5 | 16 | 480 | 72 | Yes | 0.750 | 0.173 | 0.639 | +0.727 | 0.480 | -0.193 | 1.153 |
| Study M | 21 | 42 | 1,260 | 72 | Yes | 0.823 | 0.121 | 0.685 | +0.477 | 0.888 | 0.480 | 1.297 |
| Study O | 17 | 34 | 1,187 | 72 | Yes | 0.884 | 0.067 | 0.581 | +0.505 | 0.583 | 0.079 | 1.087 |
| Study Q | 20 | 54 | 1,944 | 72 | Yes | 0.820 | 0.058 | 0.682 | -0.094 | 1.108 | 0.844 | 1.371 |
| Study R | 20 | 80 | 1,440 | 72 | Yes | 0.963 | 0.059 | 0.650 | +1.278 | 0.763 | 0.359 | 1.166 |
| Study S1 | 10 | 20 | 360 | 72 | Yes | 0.997 | 0.043 | 0.851 | +1.995 | 1.312 | -0.216 | 2.840 |
| Study S2 | 24 | 48 | 864 | 72 | Yes | 0.976 | 0.048 | 0.826 | -2.022 | 2.185 | 1.215 | 3.154 |

#### S3. Model specification

The development model was a logistic regression of character-level correctness on the standardised calibration score, fitted on character-expanded records from the development studies, with standardisation using development-study means and standard deviations only. For a session with calibration score s, the estimated accuracy is the inverse logit of a + b (s - m) / d, where m and d are the development mean and standard deviation of the score. The four numbers are given for all 18 folds in Table S12, with a worked recomputation.

Expanding each record into one row per character weights a session by the number of characters it supplied, so the quantity estimated is the accuracy of a randomly chosen character selection. That choice is stated rather than left implicit, and the reported error does not rest on it. Repeating the whole leave-one-cohort-out evaluation with each session weighted equally, and again with each participant weighted equally, gave a pooled mean absolute error of 0.096 and 0.097 against 0.098 for the character weighting. Scored on its own scale each of the three fits had a calibration slope near one, at 0.966, 0.982 and 0.977; those three slopes are on three different scales by construction, so each is comparable to one and not to the others. The three rows are written to output/expanded/estimand_comparison.csv by scripts/09_run_estimand.py.

#### S4. Uncertainty

Two uncertainty statements are reported and are not interchangeable.

The participant-cluster bootstrap used 2,000 deterministic replicates. In each replicate, development participant clusters were resampled within source study, the model was refitted, and withheld participant clusters were resampled. The resulting intervals describe uncertainty conditional on the observed set of source studies, because the same studies appear in every replicate by construction.

The same bootstrap was also used to check the between-cohort heterogeneity estimate itself, re-estimating each cohort's intercept and slope standard error from the 2,000 cluster-resampled refits and re-pooling them with the same random-effects estimator as the primary analysis. It identified 17 of the 18 cohorts, Study S1's resamples having fallen onto the separation boundary too often to leave an identified estimate, and gave tau = 0.37 for the slope and tau = 0.77 for the intercept, against 0.43 and 0.87 under the clustered sandwich (main text, Study Limitations). Those bootstrap tau values are smaller rather than larger, so this check does not itself confirm that the clustered sandwich was inflating tau. The bootstrap's own 95% interval for an unrepresented cohort runs from 0.197 to 1.861 for the slope and from -1.739 to 1.677 for the intercept, narrower than but consistent in span with the clustered-sandwich intervals reported in the main text.

The study-level summary treats the source study as the unit of replication. The withheld-cohort estimates were summarised by their mean and between-study standard deviation, with a t-distributed interval for the mean on k - 1 degrees of freedom and a prediction interval for an unrepresented cohort computed as the mean plus or minus t times the between-study standard deviation times the square root of one plus one over k.

**Table S3. Study-level summaries.** The calibration intercept and slope rows are the uncorrected summary of the 18 cohort estimates, which counts each cohort's sampling error as though it were between-cohort variation. For those two quantities the main text reports the random-effects estimate tau instead, and the intervals here should not be read in its place. The mean absolute error row is pooled on the log scale because the quantity cannot be negative; its mean and both intervals are reported back on the original scale, but its between-cohort SD is reported on the log scale, consistent with the main text. The reported mean for that row is the geometric mean; the arithmetic mean of the 18 cohort-level errors was 0.101.

| Quantity | Mean across cohorts | Between-cohort SD | 95% interval for the mean | 95% interval for an unrepresented cohort |
| --- | --- | --- | --- | --- |
| Mean absolute error (geometric mean) | 0.090 | 0.477 | 0.071 to 0.115 | 0.032 to 0.254 |
| Brier skill score | 0.174 | 0.217 | 0.066 to 0.282 | -0.296 to 0.644 |
| Character-weighted AUC | 0.713 | 0.102 | 0.662 to 0.764 | 0.491 to 0.935 |
| Calibration intercept | -0.008 | 1.175 | -0.592 to 0.576 | -2.555 to 2.539 |
| Calibration slope | 1.100 | 0.560 | 0.822 to 1.379 | -0.113 to 2.313 |

Reading the gap between the clustered sandwich (0.43/0.87) and the bootstrap (0.37/0.77) as evidence about the variance-estimation method assumes the two are pooling the same cohorts, and they are not: the bootstrap's 0.37/0.77 excludes Study S1, one of the more extreme calibration intercepts in the archive, while the clustered sandwich's 0.43/0.87 includes it. Part of the gap could therefore be Study S1's absence rather than the estimator. To separate the two, the clustered-sandwich pooling was repeated on the identical 17 cohorts the bootstrap identified, using the same random-effects estimator with Study S1 excluded (random_effects_matched_cohorts, src/bigp3_als/heterogeneity.py). That matched-cohort clustered-sandwich tau was 0.44 for the slope and 0.87 for the intercept, essentially unchanged from the all-18 values of 0.43 and 0.87 and, if anything, fractionally farther from the bootstrap's own 0.37 and 0.77 than the all-18 values were. Excluding Study S1 from the clustered-sandwich pooling therefore does not on its own reproduce the bootstrap's smaller tau, so most of the gap between the two methods reflects the variance-estimation method itself rather than Study S1's exclusion.

**Table S4. Per-cohort bootstrap replicate diagnostics.** Successful replicates are those of the 2,000 participant-cluster resamples that survived every guard in _bootstrap_standard_errors: a resampled design matrix with full column rank, a GLM refit that converges without raising, and fitted probabilities that stay clear of the 0/1 boundary by more than FITTED_BOUNDARY. A cohort is reported as identified under the bootstrap method only when at least half of the 2,000 replicates (1,000) survive; the standard error is then the sample standard deviation of exactly that surviving set, with no further filtering. Study S1 is the only cohort below that threshold.

| Cohort | Successful replicates (of 2,000) | Identified |
| --- | --- | --- |
| Study A | 2,000 | yes |
| Study B | 2,000 | yes |
| Study D | 2,000 | yes |
| Study E | 1,999 | yes |
| Study F | 2,000 | yes |
| Study G | 2,000 | yes |
| Study H | 2,000 | yes |
| Study I | 2,000 | yes |
| Study J | 2,000 | yes |
| Study K | 1,997 | yes |
| Study L | 2,000 | yes |
| Study M | 2,000 | yes |
| Study N | 2,000 | yes |
| Study O | 2,000 | yes |
| Study Q | 2,000 | yes |
| Study R | 2,000 | yes |
| Study S1 | 843 | no |
| Study S2 | 2,000 | yes |

Every cohort but Study S1 loses at most 3 of 2,000 replicates to the guards; Study S1 loses 1,157, comfortably past the 1,000-replicate threshold, which is why it alone is reported as not identified under the bootstrap method while every other cohort is estimated from an essentially complete set of replicates.

A penalized calibration model, such as Firth logistic regression, is a preferable rather than required alternative for a future analysis: refitting every bootstrap replicate with a penalized estimator would let Study S1's replicates stay identified during resampling instead of being discarded. That change would replace the estimator used inside _bootstrap_standard_errors for every replicate of every cohort, not only Study S1's, and is a materially larger undertaking than the matched-cohort comparison above. It was not pursued in this analysis and is recorded here as a deferred alternative rather than implemented.

A further check addresses a different limitation of the bootstrap above: it resamples each held-out cohort's participants independently within its own fold, so it cannot see that any two leave-one-study-out folds share up to 16 of their 17 development cohorts. joint_bootstrap_fold_covariance (src/bigp3_als/validation.py) instead draws one joint resample of every cohort's participants per replicate, using the same RANDOM_SEED and BOOTSTRAP_REPETITIONS (2,000) as the rest of this pipeline, and refits all 18 leave-one-study-out folds from that single resampled dataset, so a cohort that sits in several folds' development sets contributes the identical resampled draw to every one of them rather than an independent draw per fold. Every cohort but Study S1 produced a finite, guarded calibration fit in at least 1,997 of the 2,000 replicates. Study S1 produced one in only 826 of 2,000, below the 1,000-replicate threshold this method uses to call a cohort identified, consistent with its position near the separation boundary already documented above (Table S4) and in the main text (Study Limitations). Unlike the bootstrap in Table S4, which drops a non-identified cohort from pooling entirely, this joint bootstrap's headline figures in the Discussion (0.79 for the slope, 1.62 for the intercept) include Study S1's 826 surviving replicates alongside the other 17 cohorts' in every replicate where Study S1's fold happened to converge. The two bootstraps therefore treat a non-identified cohort differently by design, not because the pooling rule changed between them: the per-cohort bootstrap reports a single pooled tau for which an unidentified cohort has no defensible sampling distribution to contribute, while the joint bootstrap's replicate-level spread is defined over whichever folds converge in that replicate and does not require every cohort to converge in every replicate.

The empirical covariance this joint bootstrap produces (output/expanded/joint_bootstrap_covariance.csv, joint_bootstrap_correlation.csv) is not spread evenly across the 153 possible cohort pairs. Table S5 lists the eight strongest off-diagonal correlations between cohorts' held-out calibration intercepts. Four cohorts, Study F, Study J, Study L and Study Q, appear in all eight pairs, six of which pair two of them against each other, so a small subset of cohorts recurs disproportionately rather than every pair contributing comparably. Cohort size is a partial explanation: Study F and Study L are two of the smallest cohorts in the archive by participant count, at 10 and 11 of 271 (Table 2), consistent with a small cohort's resampled composition varying more from replicate to replicate and that variation propagating into every fold whose development set includes it. Study J and Study Q have 20 participants each, however, so cohort size alone does not account for the full pattern, and no single mechanism is asserted beyond the shared-development correlation this bootstrap was built to capture.

**Table S5. Strongest off-diagonal correlations between cohorts' held-out calibration intercepts under the joint bootstrap.** Correlations are Pearson correlations of the 2,000 per-replicate intercept estimates for the two named cohorts' leave-one-study-out folds. The full 36-by-36 term-by-term matrix, including the corresponding slope correlations, is output/expanded/joint_bootstrap_correlation.csv.

| Cohort pair | Correlation |
| --- | --- |
| Study L and Study Q | 0.361 |
| Study F and Study J | -0.311 |
| Study J and Study Q | -0.298 |
| Study F and Study L | 0.276 |
| Study F and Study Q | 0.267 |
| Study J and Study L | -0.263 |
| Study J and Study M | -0.262 |
| Study A and Study L | 0.248 |

#### S5. Association at three levels

**Table S6. Association between calibration-derived decoder discriminability and observed accuracy, by level and by cohort.** Pearson intervals are two-sided 95% intervals on the Fisher z scale. The two preceding-session rows order a participant's sessions by the lexical order of their identifiers, which the archive assigns sequentially within a participant; the de-identified timestamps cannot confirm that ordering, so those two rows rest on the assumption that identifier order matches recording order.

*By level.*

| Level | n | Pearson r | 95% CI | Spearman rho |
| --- | --- | --- | --- | --- |
| Sessions, pooled | 410 | 0.716 | 0.665 to 0.760 | 0.718 |
| Sessions, centred within cohort | 410 | 0.677 | 0.621 to 0.726 | 0.646 |
| Participants | 271 | 0.714 | 0.650 to 0.768 | 0.756 |
| Preceding session to later session | 139 pairs | 0.513 | 0.379 to 0.626 | 0.462 |
| Same session, reference for the row above | 139 pairs | 0.728 | 0.639 to 0.798 | 0.651 |

*By cohort.* ALS cohorts are listed first. The unit is the participant-session, so n is smaller than the record count in Table S3.

| Cohort | Sessions | Pearson r | 95% CI |
| --- | --- | --- | --- |
| Study B | 56 | 0.678 | 0.505 to 0.798 |
| Study F | 30 | 0.924 | 0.846 to 0.964 |
| Study L | 11 | 0.922 | 0.719 to 0.980 |
| Study N | 16 | 0.823 | 0.553 to 0.937 |
| Study A | 13 | 0.928 | 0.771 to 0.979 |
| Study D | 17 | 0.610 | 0.183 to 0.843 |
| Study E | 8 | 0.683 | -0.042 to 0.937 |
| Study G | 20 | 0.757 | 0.473 to 0.899 |
| Study H | 16 | 0.190 | -0.338 to 0.627 |
| Study I | 13 | 0.513 | -0.053 to 0.830 |
| Study J | 20 | 0.359 | -0.099 to 0.692 |
| Study K | 8 | 0.401 | -0.424 to 0.862 |
| Study M | 21 | 0.660 | 0.320 to 0.850 |
| Study O | 34 | 0.364 | 0.030 to 0.625 |
| Study Q | 53 | 0.773 | 0.636 to 0.863 |
| Study R | 40 | 0.425 | 0.131 to 0.651 |
| Study S1 | 10 | 0.316 | -0.392 to 0.789 |
| Study S2 | 24 | 0.609 | 0.273 to 0.813 |

The point estimate was positive in all 18 contributing cohorts. Within-cohort Pearson r ranged from 0.190 to 0.928 with a median of 0.635, and the 95% interval included zero in 6 of the 18: Study E, Study H, Study I, Study J, Study K and Study S1. All six carry 20 sessions or fewer, so the imprecision is largely a matter of cohort size rather than of a different relationship.

Session accuracy clustered within participant with an intraclass correlation of 0.412 across 410 sessions in 271 participants, giving approximately 338 effective independent sessions. The predictor is constant within a session, so the 739 session-condition records carry 410 distinct predictor values.

**Figure S1. Calibration-derived decoder discriminability against observed online accuracy, by cohort type.** Each point is one participant-session-condition record, sized by the number of eligible selections. No line is fitted to either group. Cohort type does not vary between the records of a cohort, so the comparison of the ALS cohorts against the rest is made on the 18 cohort-specific slopes, reported in the main text and drawn in Figure 1; fitting it across these records would treat a study-level attribute as though it had been measured once per record. The band of points at 1.0 is the ceiling described in the main text.

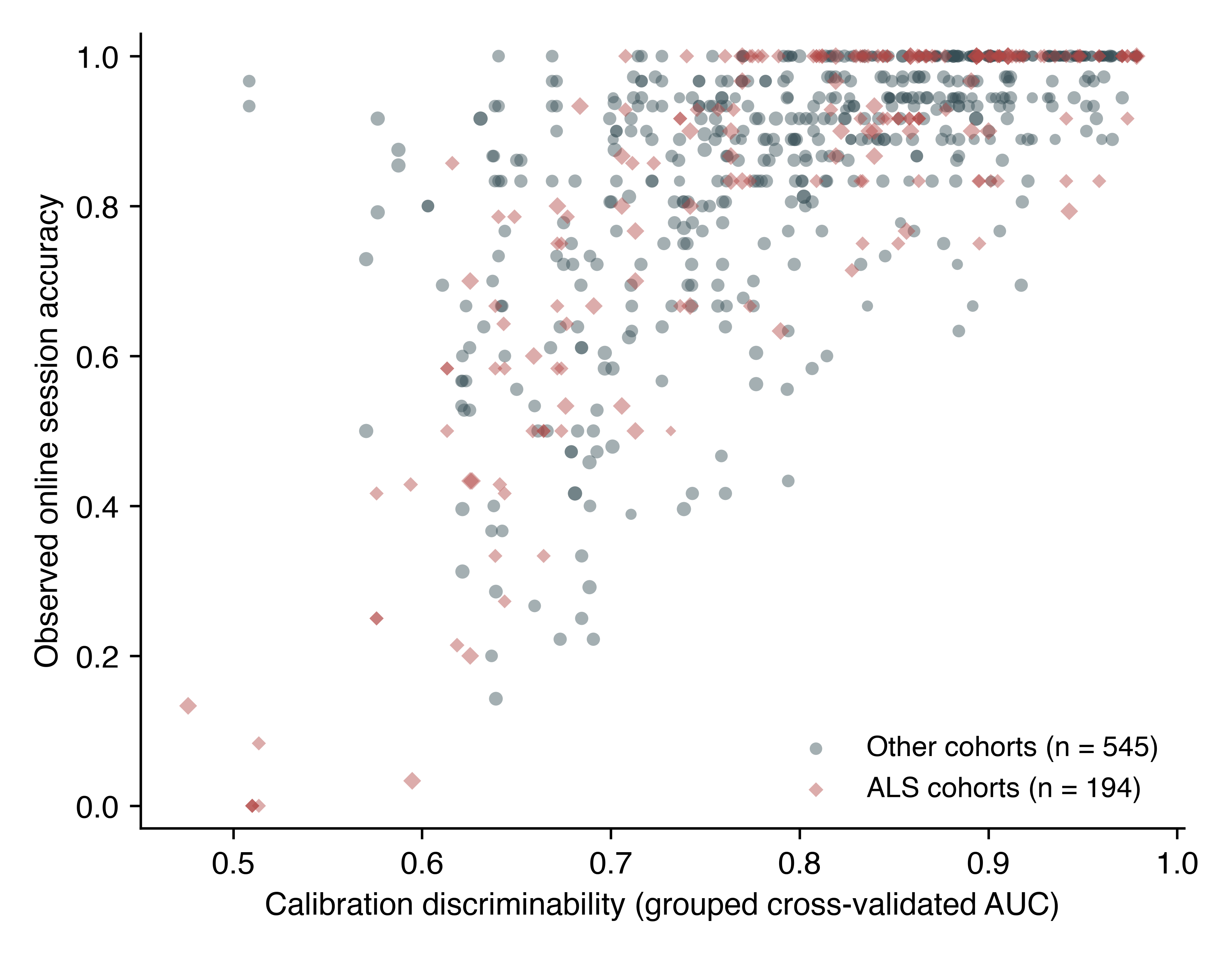

#### S6. Sensitivity analyses

**Table S7. Sensitivity analyses.** The four analyses named in the Methods, restriction to cohorts with meaningful outcome variance, exclusion of records with high artifact rejection, exclusion of small-denominator records, and leave-two-studies-out development, were listed before any of them was run on the widened cohort; none was registered. The primary row and the two cohort-subset rows are shown beside them for comparison. Each analysis re-runs the complete withheld-cohort procedure on the retained data. In the last row every pair of cohorts is withheld together, so development runs on 16 cohorts rather than the 17 of the primary analysis; each cohort appears in 17 of the 153 pairs and its 17 estimates are averaged into a single value before the same across-cohort pooling is applied, because a pair shares a cohort with 32 other pairs and the pairs are not independent units. Every row therefore summarises 18 cohort-level values, or fewer where the analysis drops cohorts, and the columns carry the same meaning throughout. The mean absolute error column (geometric mean) is the mean of the cohort-level errors, not the pooled error over records, and the two are different quantities: the ALS row reads 0.092 here, the mean of that subgroup's four cohort errors, where the main text reports 0.091 for the same analysis pooled over all of its withheld records. The mean absolute error column is pooled on the log scale because the quantity cannot be negative, as in the main text and Table S3, so its between-cohort SD is on that scale. Table S9 draws the same distinction between its pooled and cohort-mean columns.

| Analysis | Cohorts | Mean absolute error | Between-cohort SD | Interval for an unrepresented cohort | Calibration-slope SD |
| --- | --- | --- | --- | --- | --- |
| Primary, all contributing cohorts | 18 | 0.090 | 0.477 | 0.032 to 0.254 | 0.560 |
| Cohorts with outcome SD at least 0.10 | 12 | 0.117 | 0.371 | 0.050 to 0.274 | 0.601 |
| Records with artifact rejection at most 20% | 18 | 0.081 | 0.422 | 0.032 to 0.201 | 0.465 |
| Records with at least 10 eligible selections | 18 | 0.090 | 0.477 | 0.032 to 0.254 | 0.560 |
| Cohorts without a documented ALS population | 14 | 0.089 | 0.502 | 0.029 to 0.274 | 0.663 |
| ALS cohorts, originally planned subgroup | 4 | 0.092 | 0.177 | 0.049 to 0.173 | 0.223 |
| Leave-two-studies-out development, 153 splits | 18 | 0.091 | 0.477 | 0.032 to 0.255 | 0.560 |

Artifact rejection had a median of 0.002 across records, but 110 of 739 records exceeded 20% and the maximum was 0.97. The analysis restricted to records at or below 20% rejection is the only one in which both estimation error and calibration-slope variability improved.

Observed session-condition accuracy was at 100% in 228 of 739 records (30.9%). Three cohorts had mean accuracy at or above 0.96, where there is little variation to estimate.

The leave-two-studies-out row is indistinguishable from the primary row at the precision printed above, and that is the result rather than a rounding artefact. At four decimal places the mean absolute error is 0.0906 against 0.0905, its between-cohort standard deviation 0.4771 against 0.4771, and the uncorrected calibration-slope standard deviation 0.5599 against 0.5596. Removing one cohort from a development set of 17 neither raises the estimation error nor widens the between-cohort spread, so the transportability failure is not a consequence of the development set being too small at this scale. It says nothing about development sets much smaller than 16, which this design cannot examine while still leaving enough cohorts to withhold.

##### Protocol-descriptor moderator robustness

The main-text Results (Protocol Descriptors as Moderators) reports the headline association and points here for the full robustness detail.

**No other descriptor was associated with a share distinguishable from zero, including the archive's own documented grid size and its documented use of a checkerboard-variant paradigm, alongside both empirical proxies for matrix size.** The largest target index and the number of distinct characters copied bound the alphabet actually spelled from below, and both were flat: 0.003 per standard deviation (p = 0.98) and -0.019 (p = 0.89), each leaving the residual between-cohort variance at or above its unmoderated value. The archive's own descriptor documents an intended grid size (36 or 72 cells) and a stimulus paradigm per study; both were tested directly and were likewise flat: grid size at -0.061 per standard deviation (95% CI -0.343 to 0.222, p = 0.65, Holm p = 1.00) and the checkerboard-paradigm indicator at 0.176 (95% CI -0.072 to 0.423, p = 0.15, Holm p = 1.00), the latter nominally accounting for 15% of the between-cohort variance before correction. Neither test has meaningful power at this sample size: grid size takes only two values among these 18 cohorts and is confounded with ALS-cohort status (three of the four ALS cohorts use the 36-cell grid, the fourth the 72-cell grid), and the checkerboard indicator separates only 2 non-checkerboard cohorts from 16 checkerboard cohorts. The number of stimulus conditions was likewise flat (p = 0.84). Two descriptors had point estimates that were not negligible but were indistinguishable from zero once the ten tests were corrected for: the median number of selections per record at 12% (-0.215 per standard deviation, 95% CI -0.478 to 0.049, Holm p = 0.83) and the fraction of records at ceiling at 11% (0.289, 95% CI -0.022 to 0.599, Holm p = 0.60). The ceiling fraction is reported as a diagnostic rather than as a protocol difference, because it governs whether a cohort's slope is identified at all, and mean accuracy and its between-record standard deviation are summaries of the same outcomes the slope is fitted to. Entered beside the selection interval, both of the non-negligible descriptors collapse while the interval holds: with the ceiling fraction the interval is 0.397 (p = 0.011) against 0.015 (p = 0.93), and with the median number of selections it is 0.404 (p = 0.009) against 0.001 (p = 0.99). Each is a weak proxy for the interval rather than the reverse. Entered all at once the joint tests are diluted by the null moderators and neither reaches significance, the seven protocol descriptors together accounting for 42% of the between-cohort variance (F = 1.87 on 7 and 10 degrees of freedom, p = 0.18) and all ten for 14% (p = 0.39); the interval keeps its own coefficient in the seven-descriptor fit at 0.598 (p = 0.030), and loses it among all ten (p = 0.10), where 10 moderators are fitted on 18 cohorts.

**The interval is not a fully exogenous descriptor, and the result should be read with that.** Where the stopping rule was data-dependent, a session in which evidence accrued quickly ended sooner, so part of the interval is a consequence of the recording rather than a setting fixed before it. Splitting the cohorts on whether their within-recording interval never varied separates the two cases: in the 8 cohorts with a fixed interval the coefficient was 0.350 per standard deviation with a share of 48% (p = 0.14), and in the 10 with a variable interval 0.243 with a share of 55% (p = 0.046). Neither subgroup has the power of the pooled fit and the agreement in sign is what the split supports. The interval is built entirely from selection timestamps and never uses whether a selection was correct, unlike three of the ten descriptors. The verdict does not depend on how the correction family is defined: the interval's Holm p value is 0.011 within the seven protocol descriptors, 0.015 within all ten, and 0.029 when the meta-regression and the rank correlation are pooled into one family of twenty, while the ceiling fraction remains null under every definition, including 0.20 within the three outcome summaries alone. These are the same 18 cohort slopes that serve as the outcome in the preceding subsection, where cohort type was corrected separately.

#### S7. The two no-predictor benchmarks, per cohort

Skill in the main text is computed against the development-mean benchmark, which estimates every withheld record at the development-set mean accuracy. A second benchmark estimates every withheld record at that cohort's own mean, which no deployment would know. The two are different comparisons and give different counts of cohorts with negative skill, so both are given here per cohort.

Pooled, the own-mean benchmark is the harder of the two, at 0.123 against 0.146. That ordering does not hold cohort by cohort. Mean absolute error is minimised by the median rather than the mean, so a cohort's own mean is not guaranteed to beat any other constant, and it is in fact the easier target in Study A, Study F, Study K, Study L, Study M, Study N and Study Q.

Skill against the development mean is negative in one cohort, Study H. Skill against the cohort's own mean is negative in six: Study E, Study H, Study J, Study R, Study S1 and Study S2. Study S1's value is an artefact of a near-zero denominator rather than a comparable failure, because its own-mean benchmark errs by 0.005; it is reported for completeness and should not be read on the same scale as the others.

**Table S8. Model error against both no-predictor benchmarks, by withheld cohort.** ALS cohorts are listed first. Skill is one minus the ratio of the model's mean absolute error to that benchmark's.

| Cohort | Mean observed accuracy | Model MAE | Development-mean benchmark MAE | Skill vs development mean | Own-mean benchmark MAE | Skill vs own mean |
| --- | --- | --- | --- | --- | --- | --- |
| Study B | 0.899 | 0.108 | 0.158 | 0.317 | 0.132 | 0.185 |
| Study F | 0.778 | 0.101 | 0.203 | 0.502 | 0.221 | 0.543 |
| Study L | 0.839 | 0.084 | 0.137 | 0.383 | 0.140 | 0.397 |
| Study N | 0.694 | 0.124 | 0.226 | 0.448 | 0.228 | 0.454 |
| Study A | 0.786 | 0.126 | 0.173 | 0.271 | 0.178 | 0.291 |
| Study D | 0.896 | 0.056 | 0.092 | 0.394 | 0.071 | 0.211 |
| Study E | 0.921 | 0.063 | 0.079 | 0.205 | 0.049 | -0.286 |
| Study G | 0.886 | 0.066 | 0.113 | 0.417 | 0.099 | 0.336 |
| Study H | 0.877 | 0.153 | 0.121 | -0.268 | 0.109 | -0.407 |
| Study I | 0.658 | 0.172 | 0.226 | 0.236 | 0.208 | 0.171 |
| Study J | 0.708 | 0.186 | 0.187 | 0.003 | 0.185 | -0.008 |
| Study K | 0.750 | 0.173 | 0.193 | 0.104 | 0.206 | 0.160 |
| Study M | 0.823 | 0.121 | 0.134 | 0.093 | 0.138 | 0.118 |
| Study O | 0.884 | 0.067 | 0.081 | 0.181 | 0.070 | 0.046 |
| Study Q | 0.820 | 0.058 | 0.096 | 0.394 | 0.099 | 0.409 |
| Study R | 0.963 | 0.059 | 0.131 | 0.547 | 0.047 | -0.263 |
| Study S1 | 0.997 | 0.043 | 0.151 | 0.717 | 0.005 | -7.090 |
| Study S2 | 0.976 | 0.048 | 0.141 | 0.657 | 0.035 | -0.363 |
| Pooled | 0.851 | 0.098 | 0.146 | 0.327 | 0.123 | 0.203 |

#### S8. Comparator predictors

Comparator scores were computed from the identical calibration epochs and evaluated under the identical withheld-cohort protocol: regularised linear discriminant analysis area under the curve, grouped cross-validated classification accuracy, mean target-minus-non-target amplitude at Pz between 250 and 500 ms, the same contrast averaged over six posterior channels, and the maximum posterior signed r-squared. An exploratory specification adds ALSFRS-R to the primary score.

**Table S9. Withheld-cohort performance of every predictor named in the Methods.** Pooled values are computed over all withheld predictions, as in the main text. The last five columns summarise the cohort-level results and are the transportability quantities: the geometric mean across cohorts is the quantity Table S7 reports as mean absolute error and is what the interval is centred on, the interval is for a cohort not represented in the archive, and the calibration-slope spread is uncorrected, as in Table S7. Pooled MAE and the mean across cohorts are different quantities and the first is not the centre of the interval. The mean across cohorts is pooled on the log scale because the quantity cannot be negative, as in Table S3 and Table S7, so the between-cohort MAE SD is on that scale. Rows are ordered by pooled error within each role. **The last two rows rest on 3 cohorts and 138 records, not 18 and 739, so their apparently lower error is a property of that restricted set and is not comparable with the rows above.**

| Predictor | Role | Cohorts | Records | Pooled MAE | Brier skill | AUC | Pooled slope | Geometric mean across cohorts | Between-cohort SD of log(MAE) | Interval for an unrepresented cohort | Calibration-slope SD | Calibration-slope range |
| --- | --- | --- | --- | --- | --- | --- | --- | --- | --- | --- | --- | --- |
| Calibration-derived decoder discriminability | Primary | 18 | 739 | 0.098 | 0.110 | 0.748 | 0.967 | 0.090 | 0.477 | 0.032 to 0.254 | 0.560 | 0.185 to 2.185 |
| Shrinkage linear discriminant AUC | Comparator | 18 | 739 | 0.100 | 0.105 | 0.745 | 0.960 | 0.092 | 0.459 | 0.034 to 0.249 | 0.562 | 0.112 to 2.077 |
| Calibration classification accuracy | Comparator | 18 | 739 | 0.106 | 0.105 | 0.734 | 0.930 | 0.097 | 0.476 | 0.034 to 0.271 | 1.104 | 0.051 to 4.018 |
| Maximum posterior signed r-squared | Comparator | 18 | 739 | 0.134 | 0.037 | 0.656 | 0.643 | 0.129 | 0.312 | 0.066 to 0.254 | 2.209 | -0.467 to 7.297 |
| Posterior target-minus-non-target amplitude | Comparator | 18 | 739 | 0.148 | 0.003 | 0.517 | 0.275 | 0.143 | 0.277 | 0.079 to 0.261 | 2.459 | -4.745 to 6.387 |
| Pz target-minus-non-target amplitude | Comparator | 18 | 739 | 0.150 | 0.001 | 0.465 | -0.238 | 0.145 | 0.272 | 0.081 to 0.262 | 4.490 | -8.235 to 9.007 |
| Primary score plus ALSFRS-R | Exploratory | 3 | 138 | 0.085 | 0.313 | 0.834 | 0.982 | 0.089 | 0.098 | 0.055 to 0.145 | 0.293 | 0.771 to 1.336 |
| Primary score, same restricted records | Exploratory reference | 3 | 138 | 0.085 | 0.315 | 0.833 | 0.995 | 0.089 | 0.114 | 0.051 to 0.158 | 0.303 | 0.765 to 1.355 |

No confidence intervals from the participant bootstrap are reported in this table. The primary predictor's intervals are given in the main text from 2,000 replicates, and rerunning that bootstrap at a smaller replicate count for a seven-specification sweep would put a second, slightly different interval for the same primary quantity into the same paper. The uncertainty reported instead is the between-cohort spread, which requires no resampling and is the quantity the transportability question turns on. The primary row reproduces the pooled values reported in the main text on every column shown here, which is the consistency check this table can offer.

The regularised linear discriminant computed from the identical features agrees with the primary score to within 0.007 on most summary columns, the largest of those being the pooled calibration slope at 0.960 against 0.967 and the between-cohort spread of that slope at 0.562 against 0.560. Two columns separate by more than that: the per-cohort slope range, 0.112 to 2.077 against 0.185 to 2.185, and the between-cohort MAE SD, which is computed on the log scale and reads 0.459 against 0.477, a gap of 0.018. The primary result is therefore a property of how separable the calibration data are and not of the classifier family used to measure that. It carries no further implication, because the two classifiers read the same features and so cannot separate any question about how those features were built.

Scoring the same classifier by classification accuracy rather than by area under the curve costs little in pooled error, 0.106 against 0.098, but doubles the spread of the calibration slope, to 1.104 across a range of 0.051 to 4.018. A thresholded summary of the same calibration data transports worse than the ranking one.

The three summaries computed without fitting a classifier are much weaker. The maximum posterior signed r-squared reaches an area under the curve of 0.656, and neither amplitude contrast separates outcomes in a cohort it was not developed on, at 0.517 and 0.465 with Brier skill of 0.003 and 0.001. Their pooled calibration slopes of 0.275 and -0.238, and per-cohort slope ranges of -4.7 to 6.4 and -8.2 to 9.0, are what the calibration parameters look like when a predictor carries almost no transportable signal. They are reported so that the primary score's own spread is read against that scale rather than in isolation.

The exploratory ALSFRS-R specification is restricted to the 138 records in 3 cohorts that carry an observed value, so its numbers are not comparable with the rows above. Against the primary score run on those same 138 records it changes nothing: pooled error 0.085 against 0.085, area under the curve 0.834 against 0.833, Brier skill 0.313 against 0.315, and calibration-slope spread 0.293 against 0.303. The better appearance of both rows relative to the full archive belongs to the restriction and not to ALSFRS-R. At 3 cohorts the interval for an unrepresented cohort rests on two degrees of freedom and carries no transportability claim.

#### S9. Predictor reliability and disattenuated heterogeneity

The calibration score is not measured equally well in every cohort. The median number of calibration files per session ranges from 3 to 15 across cohorts, the median number of surviving calibration epochs from 1,479 to 12,910, and the standard error of the score itself from 0.008 to 0.031, a fourfold spread, tabulated per cohort in predictor_precision.csv. The three cohorts with the lowest calibration slopes, Study H at 0.185, Study J at 0.326 and Study K at 0.480, are also the three whose score was least precisely estimated, at standard errors of 0.031, 0.023 and 0.026 against a median of 0.013. Measurement error in a predictor attenuates a fitted slope toward zero, so unequal measurement error is a competing explanation for part of the between-cohort variation in calibration slope, and it is not one that collecting more cohorts would remove. This section quantifies it rather than leaving it acknowledged.

The quantity that answers the objection is the reliability ratio, lambda: the share of the observed between-session variance in a cohort's calibration score that is true signal rather than measurement error, computed as one minus the mean per-session error variance divided by the between-session variance of the score within that cohort. Per-session error variance is the Hanley and McNeil variance of an area under the curve. Classical attenuation theory gives the corrected slope as the observed slope divided by lambda, and by the delta method its standard error is divided by the same factor, so that measurement error identical in every cohort leaves the heterogeneity statistic unchanged. The 18 corrected estimates were then re-pooled with the same random-effects estimator used for the headline result, which makes the two directly comparable.

**Table S10. Reliability of the calibration score and the disattenuated calibration slope, by cohort.** ALS cohorts are listed first. Sessions are the sessions that entered that cohort's fit. Reliability is the share of the observed between-session variance in the score that is not measurement error; a value of 1 would mean the score was measured without error. The disattenuated slope is the observed slope divided by that cohort's reliability.

| Cohort | Sessions | Reliability | Observed slope | Disattenuated slope |
| --- | --- | --- | --- | --- |
| Study B | 56 | 0.984 | 1.578 | 1.603 |
| Study F | 30 | 0.987 | 1.418 | 1.437 |
| Study L | 11 | 0.992 | 1.775 | 1.789 |
| Study N | 16 | 0.977 | 1.145 | 1.173 |
| Study A | 13 | 0.987 | 1.999 | 2.025 |
| Study D | 17 | 0.979 | 0.839 | 0.857 |
| Study E | 8 | 0.982 | 0.782 | 0.797 |
| Study G | 20 | 0.969 | 1.399 | 1.443 |
| Study H | 16 | 0.848 | 0.185 | 0.218 |
| Study I | 13 | 0.965 | 1.044 | 1.082 |
| Study J | 20 | 0.950 | 0.326 | 0.344 |
| Study K | 8 | 0.957 | 0.480 | 0.501 |
| Study M | 21 | 0.983 | 0.888 | 0.904 |
| Study O | 34 | 0.964 | 0.583 | 0.605 |
| Study Q | 53 | 0.949 | 1.108 | 1.167 |
| Study R | 40 | 0.976 | 0.763 | 0.782 |
| Study S1 | 10 | 0.977 | 1.312 | 1.343 |
| Study S2 | 24 | 0.953 | 2.185 | 2.292 |

Reliability ran from 0.848 to 0.992 with a median of 0.976, and 16 of the 18 cohorts were at or above 0.95. Every cohort's reliability was identified; none was floored, clipped or dropped. The lowest, Study H at 0.848, is the cohort with the flattest slope and the least precisely measured score, so the correction acts most strongly exactly where the Discussion says the competing explanation is most plausible. It moves that cohort's slope from 0.185 to 0.218, which is not the region of 1 that a transportable mapping would occupy.

Correcting all 18 cohorts moved the between-cohort standard deviation of the slope from tau = 0.4319 to 0.4222 and I^2^ from 79.09 to 77.17, with the Q test at p = 3.6 x 10^-9 against p = 2.3 x 10^-10. The range of the cohort slopes widened rather than narrowed, from 0.185 to 2.185 observed to 0.218 to 2.292 disattenuated, because the cohorts with the flattest slopes are also the ones the correction lifts most and they were already the far end of the spread. Disattenuation therefore removes about 2 percentage points of I^2^ and does not change the conclusion.

That correction rests on the Hanley and McNeil standard error, which assumes the epochs behind an area under the curve are independent draws. They are not, because they come from repeated stimulus sequences within a cross-validated session, so the true error variance is larger than assumed and every reliability above is an overestimate. The stress test below asks how much larger it would have to be. Each factor multiplies the assumed error variance before the ratio is formed. The series stops at 6.5 because the identifiability limit is 6.58: past it, at least one cohort's assumed measurement error would exceed its entire observed spread in calibration area under the curve, which amounts to declaring that cohort's score pure noise, and the method has nothing left to say there.

**Table S11. Disattenuated heterogeneity of the calibration slope under increasing assumed measurement error.** The first row is the nominal Hanley and McNeil error variance and reproduces the disattenuated result above.

| Assumed error variance, multiple of nominal | Lowest cohort reliability | tau | I^2^ | Q | p |
| --- | --- | --- | --- | --- | --- |
| 1.0 | 0.848 | 0.422 | 77.2 | 74.5 | 3.6 x 10^-9 |
| 2.0 | 0.696 | 0.412 | 75.0 | 68.1 | 4.5 x 10^-8 |
| 3.0 | 0.544 | 0.402 | 72.8 | 62.4 | 4.1 x 10^-7 |
| 4.0 | 0.392 | 0.393 | 70.5 | 57.7 | 2.5 x 10^-6 |
| 5.0 | 0.240 | 0.388 | 68.5 | 54.0 | 9.8 x 10^-6 |
| 6.0 | 0.088 | 0.388 | 67.2 | 51.8 | 2.2 x 10^-5 |
| 6.5 | 0.012 | 0.392 | 66.9 | 51.4 | 2.6 x 10^-5 |

At the far end of that range the calibration score of one cohort is being treated as almost entirely measurement error, and the between-cohort standard deviation has still only fallen from 0.432 to 0.392, I^2^ is still 66.9, and the Q test has never risen above p = 2.6 x 10^-5. Measurement error in the predictor is a real contributor to the between-cohort spread and a minor one, and it does not explain the failure of the mapping to transport.

Two limits of this analysis should be read alongside it. Reliability is estimated per cohort from as few as 8 sessions, so each ratio is itself imprecise, and the correction treats lambda as known. And attenuation is the only mechanism modelled: it addresses measurement error in the predictor and says nothing about differences in protocol, population or stopping rule, which are treated separately in the moderator analysis.

#### S10. The full 18-cohort calibration-curve figure

The main-text Figure 2 shows six representative cohorts, so that the panels are legible at the size a printed page allows. This section gives the same figure for every withheld cohort.

**Figure S2. Observed against estimated session accuracy, one panel per withheld cohort, all 18 cohorts.** The main-text Figure 2 shows six representative cohorts (the four ALS cohorts and the two non-ALS extremes of the calibration-slope range); this figure shows every cohort. Each point is one of up to ten equal-count bins of the estimate within that cohort, placed at the selection-weighted mean estimate and the observed accuracy of the records in it, with point area increasing with the selections it rests on. A cohort supports at most as many bins as it has distinct estimates, and the estimate is a property of the session rather than of the record, so two of the 18 cohorts draw eight bins rather than ten. The dashed line is equality. Points above it are bins whose accuracy the mapping understated and points below are bins whose accuracy it overstated, so the panels carry the direction of the miscalibration that Figure 1 summarises as a spread. The number in each panel is observed minus estimated accuracy across that whole cohort.

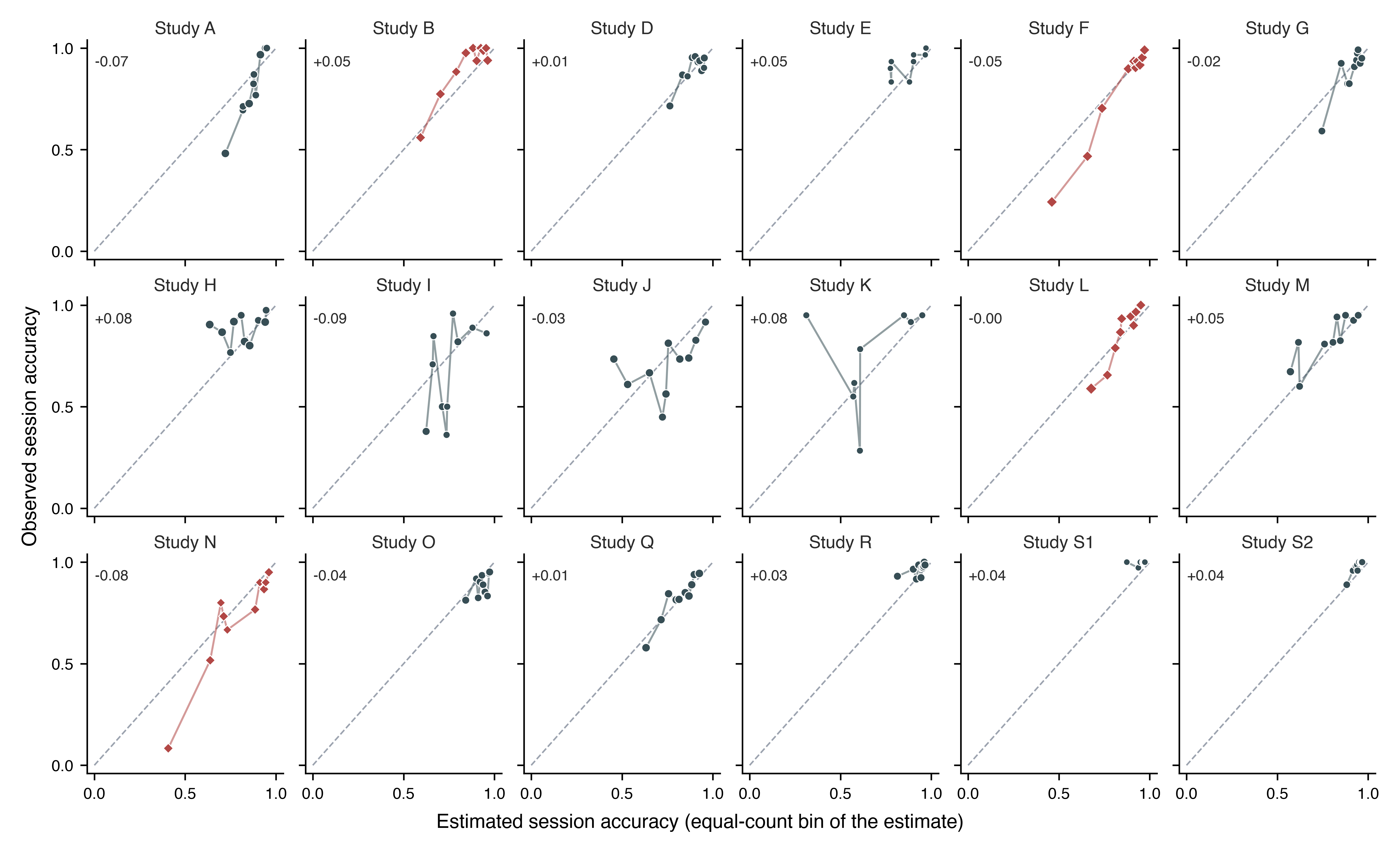

#### S11. Reproducibility

##### Recomputing any estimate

**Table S12. Fitted coefficients of every development fold.** One row per withheld cohort. The estimate for a session with calibration score s in the fold that withheld a given cohort is the inverse logit of a + b (s - m) / d, using that row's four numbers. Coefficients are on the standardised scale, so b is the change in log odds per development standard deviation of the score.

| Withheld cohort | Development records | Development selections | a | b | m | d |
| --- | --- | --- | --- | --- | --- | --- |
| Study A | 700 | 18,207 | 2.0672 | 0.9766 | 0.80553 | 0.10820 |
| Study B | 683 | 18,830 | 2.0030 | 0.9688 | 0.80476 | 0.10689 |
| Study D | 705 | 18,381 | 2.0155 | 0.9934 | 0.80354 | 0.10794 |
| Study E | 731 | 19,371 | 2.0209 | 0.9783 | 0.80449 | 0.10657 |
| Study F | 650 | 18,544 | 2.0281 | 0.8991 | 0.80508 | 0.10197 |
| Study G | 699 | 18,413 | 2.0127 | 0.9852 | 0.80218 | 0.10761 |
| Study H | 675 | 17,685 | 2.0362 | 1.0357 | 0.80886 | 0.10788 |
| Study I | 713 | 18,663 | 2.0823 | 0.9493 | 0.80817 | 0.10570 |
| Study J | 699 | 17,799 | 2.1463 | 1.0333 | 0.80942 | 0.10474 |
| Study K | 723 | 19,131 | 2.0624 | 1.0001 | 0.80719 | 0.10470 |
| Study L | 706 | 18,621 | 2.0219 | 0.9638 | 0.80551 | 0.10730 |
| Study M | 697 | 18,351 | 2.0492 | 0.9925 | 0.80866 | 0.10604 |
| Study N | 723 | 19,131 | 2.0458 | 0.9561 | 0.80577 | 0.10548 |
| Study O | 705 | 18,424 | 2.0442 | 1.0220 | 0.80138 | 0.10704 |
| Study Q | 685 | 17,667 | 2.0533 | 0.9883 | 0.80842 | 0.10811 |
| Study R | 659 | 18,171 | 1.9028 | 0.9545 | 0.79471 | 0.10654 |
| Study S1 | 719 | 19,251 | 1.9809 | 0.9553 | 0.80105 | 0.10529 |
| Study S2 | 691 | 18,747 | 1.9400 | 0.9513 | 0.79803 | 0.10621 |

*Worked example.* The first record of the cohort withheld in the Study H fold has a calibration score of 0.710714. From that row, m = 0.80886 and d = 0.10788, so the standardised score is (0.710714 - 0.80886) / 0.10788 = -0.9098. With a = 2.0362 and b = 1.0357 the log odds are 2.0362 + 1.0357 x (-0.9098) = 1.0940, and the inverse logit of that is 0.7491, which is the estimate the analysis records for that session-condition.

Recomputing all 739 held-out estimates from the printed values reproduces the frozen predictions to within 0.0001. The coefficients themselves are not refitted for this table; they are read off the fitted model at two scores, and every fold is checked by reconstructing its own predictions from the four numbers before the table is written.

##### Pipeline and outputs

The analysis pipeline executes archive validation, source metadata extraction, feedback-phase reconstruction, calibration feature extraction, withheld-cohort validation, the widened-design analyses, per-cohort calibration and heterogeneity, predictor precision and reliability with the disattenuated heterogeneity of S9, the estimand comparison, the protocol-moderator analysis, sensitivity analyses, the comparator-predictor sweep, the fold-coefficient export, the joint bootstrap of all 18 development folds, and the rendering of every figure from those frozen outputs. Frozen outputs are written to output/expanded/: study_inventory.csv, analysis_records.csv, external_validation_metrics.csv, external_validation_predictions.csv, random_effects_pooling.csv, als_subgroup_metrics.csv, transfer_to_als.csv, als_meta_regression.json, null_benchmark.csv, within_study_association.csv, participant_level_association.csv, across_session_association.csv, across_session_pairs.csv, session_clustering.json, cohort_calibration.csv, heterogeneity_summary.json, predictor_precision.csv, predictor_reliability.csv, estimand_comparison.csv, sensitivity_analyses.csv, comparator_metrics.csv, fold_coefficients.csv, protocol_covariates.csv, protocol_meta_regression.json, joint_bootstrap_covariance.csv, joint_bootstrap_correlation.csv, joint_bootstrap_replicate_diagnostics.csv, bootstrap_replicate_diagnostics.csv and joint_bootstrap_summary.json. The repository test suite contains 182 tests covering provenance, European Data Format parsing, event reconstruction, feature extraction, validation, the widened-design analyses, the promised sensitivity, comparator and fold-coefficient analyses, the session-ordering assumption behind the preceding-session analysis, and rendering. The completed TRIPOD checklist is a separate file, supplementary/tripod_checklist.md, submitted alongside this supplement rather than as a numbered section within it.

#### S12. Transparency statement

As stated in the Methods, under Ethics, the study involved no new data collection, participant contact, prospective enrolment, or intervention. It does not establish a diagnostic, prognostic, causal, or treatment effect. Character-level online selection accuracy is an operational endpoint and should not be presented as communication success, quality of life, or a clinical outcome. The source studies vary in protocol, and the archive does not permit cross-study person-level linkage. As detailed in S1, participant identifiers are scoped to each source study rather than to the archive as a whole, so cross-study participant overlap can be neither confirmed nor excluded from the archive's documentation. Four source studies carry a documented ALS population; the remaining cohorts are described as other cohorts because the documentation does not support a positive characterisation, and no participant-level clinical characteristics were available.
